# A Phase 1, Single-Center, Randomized, Double-Blind, Placebo-Controlled, Multiple-Dose Escalation Study for the Evaluation of the Safety, Tolerability, and Pharmacokinetics of Recombinant Human Plasma Gelsolin (rhu-pGSN) Following Intravenous Administration to Healthy Volunteers

**DOI:** 10.64898/2026.03.24.26348914

**Authors:** Yao Liu, Susan L Levinson, Edward Kowalik, Jeremy Pronchik, Lester Kobzik, Mark J DiNubile

## Abstract

**Background:** Plasma gelsolin (pGSN) is a non-immunosuppressive anti-inflammatory immunomodulator with demonstrated efficacy in animal models of acute lung injury. Its potential role in moderate-to-severe acute respiratory distress syndrome (ARDS) is currently under investigation.

**Methods:** We conducted a phase 1, randomized, double-blind, placebo-controlled study to evaluate the safety, tolerability, and pharmacokinetics of recombinant human pGSN (rhu-pGSN) following intravenous (IV) administration to healthy volunteers. Thirty-two participants were assigned to 4 sequentially ascending dose cohorts (6, 12, 18, 24 mg/kg of body weight) to receive five IV infusions of rhu-pGSN or saline placebo. Each cohort includes 8 subjects randomized 3:1 with rhu-pGSN or placebo. Doses were administered at 0 hours, 12 hours, 36 hours, 60 hours, and 84 hours. The primary outcome is the incidence and severity of clinical and laboratory AEs regardless of causality. Secondary outcomes include the pharmacokinetics of IV rhu-pGSN and the presence of anti-rhu-pGSN antibodies at Day 28.

**Results:** Overall, 10 subjects (41.7%) who received rhu-pGSN reported a total of 13 adverse events (AEs), and 1 subject (12.5%) who received placebo reported an AE. All AEs were mild or moderate. AEs in system organ classes that were reported by 2 or more subjects in either arm were skin and subcutaneous tissue disorders (12.5% rhu-pGSN; 0% placebo), gastrointestinal disorders (8.3% rhu-pGSN; 0% placebo), and nervous system disorders (12.5% rhu-pGSN; 12.5% placebo). No AEs by preferred term were reported by more than 1 subject in either arm. Three subjects (12.5%) experienced an AE assessed as related to study drug. No serious AEs occurred, and no AEs led to study discontinuation, dose interruption/reduction, or death. There were no apparent between-treatment differences in laboratory abnormalities, vital signs, or electrocardiogram findings.

**Conclusions:** Overall, in this study, IV rhu-pGSN (up to 24 mg/kg daily) appeared safe and well tolerated compared to placebo. The median half-life of rhu-pGSN exceeded 14 h across all dosing regimens, supporting once daily IV dosing in healthy subjects.

**Trial registration:** This study was registered with ClinicalTrials.gov on 2023-03-29 under the registration identifier NCT05789745.

## Background

Plasma gelsolin (pGSN) is a non-immunosuppressive, anti-inflammatory protein produced and secreted by virtually every cell type, and it circulates at high levels in the blood of healthy individuals (100-300 ug/ml) (1). The natural protein, and its recombinant counterpart (rhu-pGSN), are comprised of 755 amino acids and are non-glycosylated. Extensive experimental work has shown that pGSN exerts broad anti-inflammatory and host-protective effects, including scavenging extracellular actin, binding inflammatory mediators, enhancing pathogen clearance, inhibiting activation of the NLRP3 inflammasome, and promoting the transition of macrophages from a proinflammatory (M1) to an anti-inflammatory (M2) phenotype (2–8).

Circulating pGSN levels decline substantially in a wide range of acute and chronic diseases, including trauma, infections, neurologic disorders and many others (9–13). It is noteworthy that the lower the levels of plasma gelsolin, the less favorable the prognosis of acute illness becomes (9). This association has been observed in several acute lung injury (ALI) conditions, such as community-acquired pneumonia (CAP) (9), COVID-19 (14–16), ALI after cardiac bypass (17) or allogeneic stem cell transplantation (18) and ventilator weaning in critically ill patients (19).

The functional importance of pGSN in modulating inflammation is further supported by animal studies in which pGSN depletion exacerbates diseases, while rhu-pGSN administration improves outcomes across diverse models whose main common denominator is dysregulated inflammation. These include sepsis (20), arthritis (21), decompression sickness (7,22) and CO-induced neurological injury (23). Notably, rhu-pGSN has also shown consistent benefits in preclinical models of lung injury, including bacterial pneumonia (3,24), influenza (25) and hyperoxia (26).

Together, these findings provide two complementary lines of evidence supporting the therapeutic potential of rhu-pGSN administration in ARDS/ALI: (1) clinical observations linking low endogenous pGSN levels to worse outcomes, and (2) preclinical studies demonstrating improved outcome with pGSN administration in animal models of inflammatory lung injuries.

We conducted a phase-1 clinical study to evaluate the safety, tolerability and pharmacokinetics of rhu-pGSN in healthy volunteers, providing the foundation for the now ongoing phase-2 trial in moderate-to-severe ARDS (NCT05947955).

## Methods

### Objective

The primary objective of this study (NCT05789745) was to evaluate the safety and tolerability of 4 sequentially ascending doses of rhu-pGSN at 6 mg/kg, 12 mg/kg, 18 mg/kg and 24 mg/kg^a^ of actual body weight administered by intravenous (IV) infusion for 5 doses at 0, 12, 36, 60, and 84 hours to healthy subjects. Secondary objectives include characterization of the pharmacokinetic profile of rhu-pGSN, as well as investigation of the development of anti-pGSN antibodies following administration of 5 doses of rhu-pGSN to healthy subjects.

### Study Materials

Rhu-pGSN (BioAegis Therapeutics, North Brunswick, NJ) was produced in *Escherichia coli* and subsequently purified and lyophilized under good manufacturing practice (GMP) procedures. The study site received 10-ml glass vials containing lyophilized powder, which were reconstituted with 4.9 ml of sterile water to yield approximately 5.3 ml of solution at a final concentration of 40 mg/ml^a^ rhu-pGSN in a proprietary stabilizing buffer.

### Study Design and Participants

This phase I, randomized, double-blind, placebo-controlled, dose escalation, parallel design study was conducted from February 14, 2023 through May 22, 2023, by Nucleus Network at Saint Paul, Minnesota. A total of 32 healthy volunteers aged 18 to 55 years, with body weight ≤ 100 kg, and a body mass index < 30 kg/m^2^, were enrolled. The clinical study protocol and any other appropriate study-related documents were reviewed and approved by the Institutional Review Board (IRB). Written informed consent was obtained from all participants. Participants were excluded if they were pregnant or lactating women; had acute illness during the month prior to screening; required any medications during the 5 inpatient days other than acetaminophen; were hospitalized during the year prior to screening; had a history of cancer or treatment with chemotherapy or radiation therapy; had a history of transplantation; had a history of diabetes mellitus, cardiovascular disease, cerebrovascular disease, pulmonary disease, liver or kidney disease, psychiatric condition, or active or chronic infection; received blood products during the year prior to screening; underwent chronic mechanical ventilation or dialysis; had any clinically significant abnormalities in clinical chemistry, hematology, urinalysis results, vital signs, EKG, or physical examination findings as judged by the Investigator; or positive results for recreational drugs during screening. Full inclusion and exclusion criteria are listed in the Supplementary Materials.

Thirty-two healthy volunteers were assigned to 4 sequentially ascending dose cohorts (6, 12, 18, 24 mg/kg of body weight) to receive five IV infusions of rhu-pGSN or saline placebo (Fig. 1). Each cohort includes 8 subjects randomized 3:1 with rhu-pGSN or placebo (6 rhu-pGSN subjects: 2 placebo subjects). Doses were administered at 0 hours (Day 1), 12 hours (Day 1), 36 hours (Day 2), 60 hours (Day 3), and 84 hours (Day 4). Subjects stayed at the study site until after the last blood sample was taken at 108 hours (Day 5). Subjects returned for follow-up 7 days after the initiation of therapy (on Day 8) and on Day 28 for the End-of-Study (EOS) visit. After each cohort completed the Day 8 visit, a review of the safety results was conducted before the initiation of the next higher dose cohort. The entire study was 28 ± 3 days in duration.

**Fig 1.**
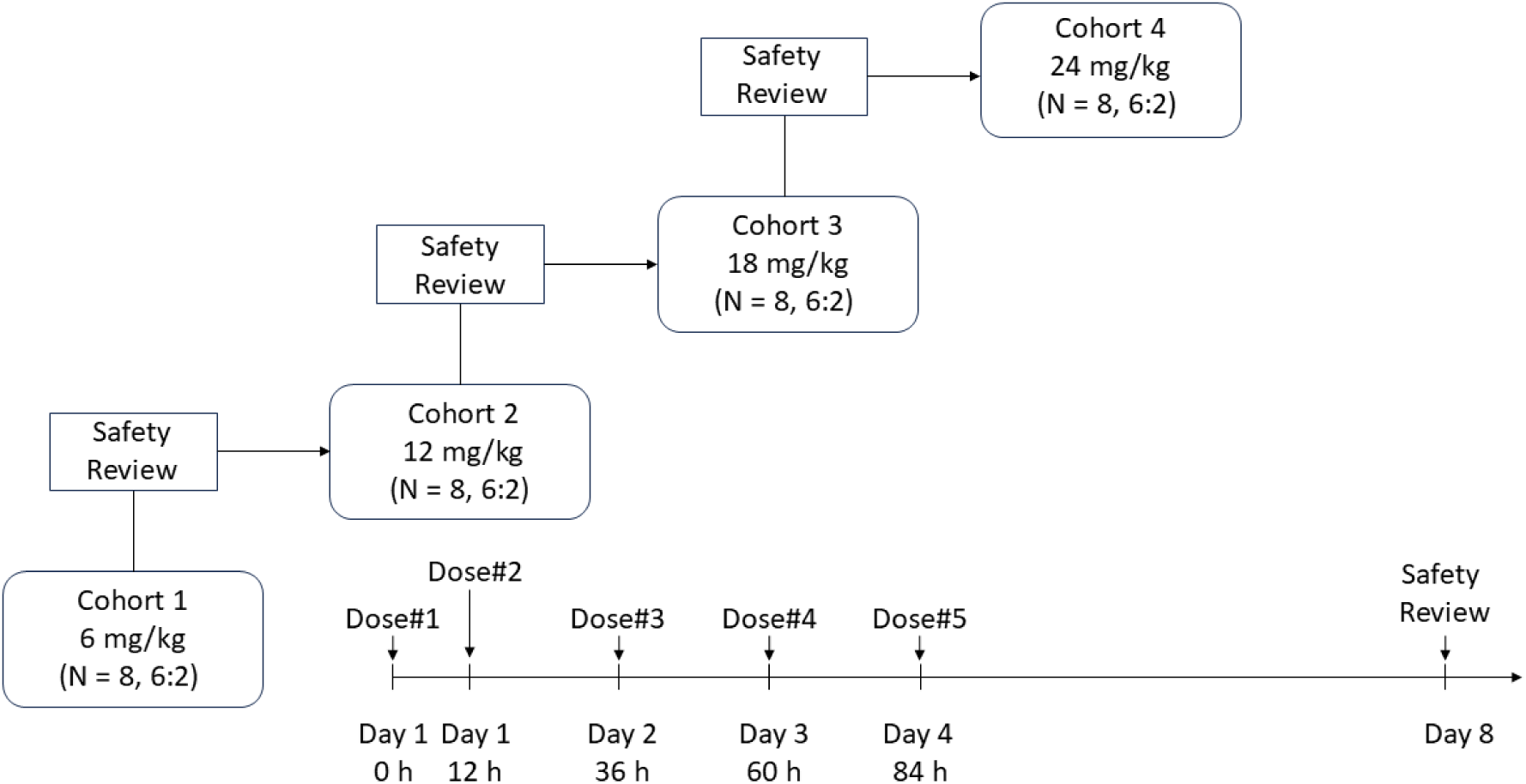
Study design and dosing regimen

### Procedures

Participants were admitted to the clinical site 1 day before study drug administration and remained there until 24 hours after the last dosing (on Day 5). Plasma samples for pharmacokinetic analysis were collected within 15 minutes before, and at 15 minutes and 1, 2, 4, 8 and 12 hours after Dose #1, #2 and #5, respectively; additional plasma samples were collected at 24 hours after Dose #2 and #5. A single plasma sample for pharmacokinetic analysis was also collected anytime on Days 8 and 28. Serum samples were collected on Day 1 before Dose #1 and on Day 28 for assessing the presence of anti-rhu-pGSN antibodies. Participants were monitored for adverse events (AEs), clinical laboratory measurements, vital signs, and electrocardiogram (EKG) measurements.

### Outcomes

AEs were coded using the Medical Dictionary for Regulatory Activities (MedDRA) version 25.1. Plasma pGSN concentrations were measured by enzyme-linked immunosorbent assay (ELISA) as previously described (27). Since the assay measured endogenous pGSN levels as well as exogenous rhu-pGSN, pharmacokinetic parameters were calculated for values above baseline by subtracting pre-dose pGSN level from each observed concentration. Baseline-adjusted concentrations that are <0 were set to 0. The pharmacokinetic parameters were calculated by Phoenix WinNonlin version 6.3 or higher (Certara, Princeton, New Jersey), which include but not limited to maximum concentration (C_max_), time to maximum concentration (T_max_), terminal half-life (t_1/2_), area under the concentration-time curve (AUC) from time 0 to 8h (AUC_0-8_), AUC from 0 to 12h (AUC_0-12_), AUC from 0 to 24h (AUC_0-24_), and AUC from 0 to infinity (AUC_0-inf_). For the assessment of anti-rhu-pGSN antibody in serum samples, a Meso Scale Diagnosistics (MSD) based electrochemiluminescent immunoassay was used. It utilizes the bivalent binding capability of anti-pGSN antibodies to form a bridging complex with biotinylated Protein G and ruthenylated pGSN to generate RLU (relative light units) for the measurement of anti-pGSN antibodies in matrix. Samples were tested in a three-tier approach. All samples were initially evaluated in a screening assay (Tier I) to detect the presence of antibody, samples with positive signals (signals above the calculated screening assay cut point value) were further evaluated in a confirmatory assay (Tier II) to determine the specificity of the reactivity. Samples confirmed positive (signals above the confirmatory assay cut point value) were evaluated in a titration assay (Tier III) to semi-quantitate the anti-rhu-pGSN antibody by reporting a titer.

### Statistical Analysis

The sample size was 32 subjects in total: 6 subjects received rhu-pGSN and 2 subjects received placebo in each of the 4 dosing cohorts. Data from all subjects who received placebo were combined into a single placebo group.

Safety was assessed in all subjects who were randomized and received at least part of one dose of study treatment (Safety Population). Pharmacokinetics were assessed in all subjects in the safety population who had at least 1 quantifiable study drug concentration post-dose (PK Population). The Per Protocol (PP) Population was to exclude all subjects in the Safety Population who had missed or mistimed doses, received the wrong dose, and/or randomly discontinued the study before the primary Day 8 visit.

Continuous variables were summarized using descriptive statistics, including the number of subjects (N) with non-missing values, mean, standard deviation (SD), median, minimum, and maximum. For PK data, geometric mean, coefficient of variation (CV%), and geometric coefficient of variation (geo CV%) values were also presented. In summary tables of categorical variables, frequency counts and percentages of subjects within each category were presented.

All statistical analysis was performed using SAS version 9.4 (SAS Institute, Cary, North Carolina).

^a^ Subsequent to conducting this study, it was determined that the extinction coefficient used historically to determine protein concentration for plasma gelsolin underreported the actual protein levels. Theoretical and experimental determination of the extinction coefficient confirms that these values are underreported by 11%. Thus, this study was performed at doses that were 11% higher than reported below. The values reported here used the previous extinction coefficient and are not adjusted.

## Results

### Participants

A total of 32 subjects were screened and all were randomized with rhu-pGSN (N=24; 6 in each of the 4 cohorts) or placebo (N=8). One subject discontinued the study due to withdrawal of consent (subject moved to another state). All other participants completed the study (Fig. 2).

**Fig 2.**
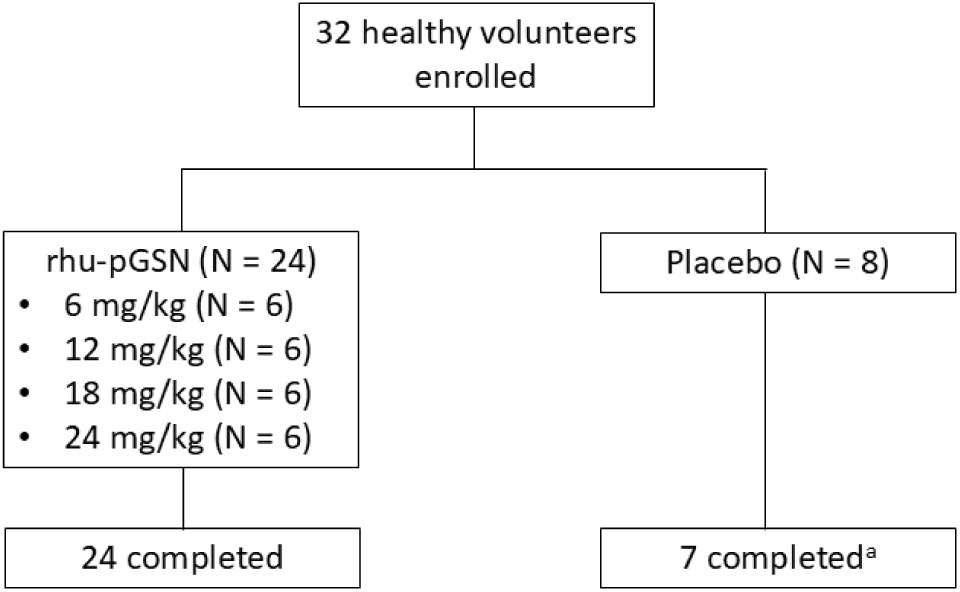
Patient disposition. ^a^One subject discontinued the study due to withdrawal of consent (subject moved to another state)

The mean (SD) age of the 32 subjects was 36.2 (11.0) years (range: 20.0 to 55.0 years; Table 1). Twenty subjects (62.5%) were male, and twelve subjects (37.5%) were female. Most subjects were White (75.0%); the remainder of the subjects were Asian or African American (12.5% each). The mean BMI (SD) was 24.7 (2.9).

**Table 1.**
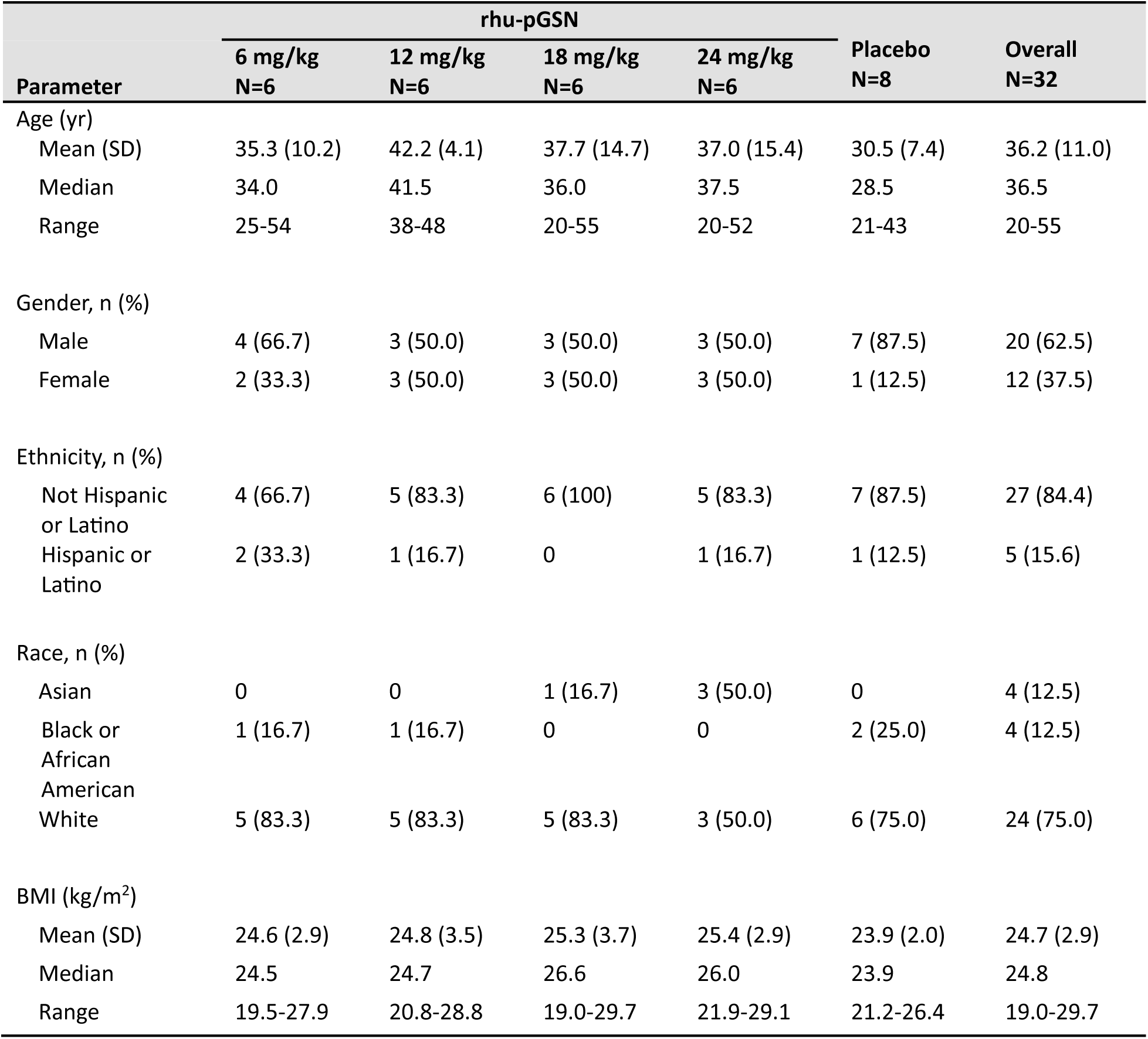
Subject demographics.

| Parameter | rhu-pGSN |  |  |  | Placebo<br>N=8 | Overall<br>N=32 |
| --- | --- | --- | --- | --- | --- | --- |
|  | 6 mg/kg<br>N=6 | 12 mg/kg<br>N=6 | 18 mg/kg<br>N=6 | 24 mg/kg<br>N=6 |  |  |
| Age (yr) |  |  |  |  |  |  |
| Mean (SD) | 35.3 (10.2) | 42.2 (4.1) | 37.7 (14.7) | 37.0 (15.4) | 30.5 (7.4) | 36.2 (11.0) |
| Median | 34.0 | 41.5 | 36.0 | 37.5 | 28.5 | 36.5 |
| Range | 25-54 | 38-48 | 20-55 | 20-52 | 21-43 | 20-55 |
| Gender, n (%) |  |  |  |  |  |  |
| Male | 4 (66.7) | 3 (50.0) | 3 (50.0) | 3 (50.0) | 7 (87.5) | 20 (62.5) |
| Female | 2 (33.3) | 3 (50.0) | 3 (50.0) | 3 (50.0) | 1 (12.5) | 12 (37.5) |
| Ethnicity, n (%) |  |  |  |  |  |  |
| Not Hispanic or Latino | 4 (66.7) | 5 (83.3) | 6 (100) | 5 (83.3) | 7 (87.5) | 27 (84.4) |
| Hispanic or Latino | 2 (33.3) | 1 (16.7) | 0 | 1 (16.7) | 1 (12.5) | 5 (15.6) |
| Race, n (%) |  |  |  |  |  |  |
| Asian | 0 | 0 | 1 (16.7) | 3 (50.0) | 0 | 4 (12.5) |
| Black or African American | 1 (16.7) | 1 (16.7) | 0 | 0 | 2 (25.0) | 4 (12.5) |
| White | 5 (83.3) | 5 (83.3) | 5 (83.3) | 3 (50.0) | 6 (75.0) | 24 (75.0) |
| BMI (kg/m<sup>2</sup>) |  |  |  |  |  |  |
| Mean (SD) | 24.6 (2.9) | 24.8 (3.5) | 25.3 (3.7) | 25.4 (2.9) | 23.9 (2.0) | 24.7 (2.9) |
| Median | 24.5 | 24.7 | 26.6 | 26.0 | 23.9 | 24.8 |
| Range | 19.5-27.9 | 20.8-28.8 | 19.0-29.7 | 21.9-29.1 | 21.2-26.4 | 19.0-29.7 |

One subject in Cohort 1 (6 mg/kg) was transgender (male to female). The gender used for study purposes was the same as that assigned on the subject’ s birth certificate (male). This ensured consistency in controlling for biological differences in physical characteristics between the sexes, such as when measuring pharmacokinetic parameters.

Demographics and baseline characteristics were generally well balanced across rhu-pGSN and placebo groups with the exception of gender; the ratio of male to female subjects in the placebo group (7:1) was higher than the ratio of male to female subjects across the rhu-pGSN groups (range: 4:2 to 3:3). Placebo recipients were younger than rhu-pGSN recipients overall.

### Safety

10 of 24 subjects (41.7%) who received rhu-pGSN reported a total of 13 AEs, 11 of which were treatment-emergent AEs (TEAEs). 1 of 8 subject (12.5%) who received placebo reported 1 AE, which was also a TEAE (Table 2). All AEs were mild or moderate. The overall proportion of subjects with AEs was higher in the 18 and 24 mg/kg dose groups (50.0%) and lower in the 6 and 12 mg/kg dose groups (33.3%). Events in any specific system organ classes occurred in ≤3 subjects in either arm (Table 3). AEs in system organ classes that were reported by 2 or more subjects in either arm were skin and subcutaneous tissue disorders (12.5% rhu-pGSN; 0% placebo), gastrointestinal disorders (8.3% rhu-pGSN; 0% placebo), and nervous system disorders (12.5% rhu-pGSN; 12.5% placebo). No AEs by preferred term were reported by more than 1 subject in either arm. Three subjects (12.5%) experienced an AE assessed as related (including definitely related, probably related and possibly related) to study drug, which were constipation, fatigue, and epigastric discomfort (Table 2, Supplementary Table 1). No AEs were serious and none led to study discontinuation, interruption or reduction of study drug, or death. There were no clear differences between the treatments over time in mean or median laboratory variables. Selected summary statistics of key hematology parameters of special interest by dose group are summarized in Supplementary Table 2. There were no between-treatment differences in vital signs, or EKG findings, either.

**Table 2.** Overview of adverse events in the Safety Population.

| Parameter | rhu-pGSN |  |  |  |  | Placebo<br>N=8 |
| --- | --- | --- | --- | --- | --- | --- |
|  | 6 mg/kg<br>N=6 | 12 mg/kg<br>N=6 | 18 mg/kg<br>N=6 | 24 mg/kg<br>N=6 | Combined<br>N=24 |  |
| No. of AEs | 2 | 3 | 5 | 3 | 13 | 1 |
| No. of Treatment-emergent AEs (TEAEs) | 1 | 3 | 5 | 2 | 11 | 1 |
| No. (%) of subjects with at least 1: |  |  |  |  |  |  |
| AE | 2 (33.3) | 2<sup>a</sup> (33.3) | 3<sup>b</sup> (50.0) | 3 (50.0) | 10 (41.7) | 1 (12.5) |
| Serious AE | 0 | 0 | 0 | 0 | 0 | 0 |
| AE leading to study discontinuation | 0 | 0 | 0 | 0 | 0 | 0 |
| AE leading to study interruption | 0 | 0 | 0 | 0 | 0 | 0 |
| AE leading to death | 0 | 0 | 0 | 0 | 0 | 0 |
| TEAE | 1 (16.7) | 2 (33.3) | 3 (50.0) | 2 (33.3) | 8 (33.3) | 1 (12.5) |
| No. (%) of subjects with AEs by NCI-CTCAE grade |  |  |  |  |  |  |
| 1 - Mild | 1 (16.7) | 2<sup>a</sup> (33.3) | 3<sup>b</sup> (50.0) | 2 (33.3) | 8 (33.3) | 1 (12.5) |
| 2 - Moderate | 1 (16.7) | 1<sup>a</sup> (16.7) | 0 | 1 (16.7) | 3 (12.5) | 0 |
| 3 - Severe | 0 | 0 | 0 | 0 | 0 | 0 |
| 4 - Life-threatening | 0 | 0 | 0 | 0 | 0 | 0 |
| 5 - Fatal | 0 | 0 | 0 | 0 | 0 | 0 |
| No. (%) of subjects with AEs by relationship to study drug |  |  |  |  |  |  |
| Definitely unrelated | 2 (33.3) | 2<sup>a</sup> (33.3) | 2<sup>b</sup> (33.3) | 2 (33.3) | 8 (33.3) | 0 |
| Probably unrelated | 0 | 0 | 1<sup>b</sup> (16.7) | 0 | 1 (4.2) | 1 (12.5) |
| Possibly related | 0 | 0 | 2<sup>b</sup> (33.3) | 1 (16.7) | 3 (12.5) | 0 |
| Probably related | 0 | 0 | 0 | 0 | 0 | 0 |
| Definitely related | 0 | 0 | 0 | 0 | 0 | 0 |
| No. (%) of subjects with AEs by relationship to study procedure |  |  |  |  |  |  |
| Definitely unrelated | 1 (16.7) | 2<sup>a</sup> (33.3) | 3<sup>b</sup> (50.0) | 3 (50.0) | 9 (37.5) | 1 (12.5) |
| Probably unrelated | 0 | 0 | 1<sup>b</sup> (16.7) | 0 | 1 (4.2) | 0 |
| Possibly related | 0 | 0 | 0 | 0 | 0 | 0 |
| Probably related | 0 | 0 | 0 | 0 | 0 | 0 |
| Definitely related | 1 (16.7) | 1<sup>a</sup> (16.7) | 0 | 0 | 2 (8.3) | 0 |

<sup>a</sup>One subject experienced two AEs: one mild and one moderate. Both AEs were deemed definitely unrelated to study drug. One AE was deemed definitely unrelated to study procedure, the other definitely related.
<sup>b</sup>One subject experienced three AEs, all of which were mild. One AE was deemed definitely unrelated to study drug, one probably unrelated, and one possibly related. Three AEs were deemed definitely unrelated to study procedure, and one probably unrelated.

**Table 3.** Summary of adverse events by System Organ Class and Preferred Term in the Safety Population.

| System Organ Class<br>Preferred Term | rhu-pGSN |  |  |  |  | Placebo<br>N=8<br>No. (%) |
| --- | --- | --- | --- | --- | --- | --- |
|  | 6 mg/kg<br>N=6 | 12 mg/kg<br>N=6 | 18 mg/kg<br>N=6 | 24 mg/kg<br>N=6 | Combined<br>N=24 |  |
|  | No. (%) | No. (%) | No. (%) | No. (%) | No. (%) |  |
| **Skin and subcutaneous tissue disorders** | 0 | 1 (16.7) | 1 (16.7) | 1 (16.7) | 3 (12.5) | 0 |
| Dermatitis | 0 | 0 | 0 | 1 (16.7) | 1 (4.2) | 0 |
| Ecchymosis | 0 | 1 | 0 | 0 | 1 (4.2) | 0 |
| Rash | 0 | 0 | 1 (16.7) | 0 | 1 (4.2) | 0 |
| **Gastrointestinal disorders** | 0 | 0 | 1 (16.7) | 1 (16.7) | 2 (8.3) | 0 |
| Constipation | 0 | 0 | 1 (16.7) | 0 | 1 (4.2) | 0 |
| Epigastric discomfort | 0 | 0 | 0 | 1 (16.7) | 1 (4.2) | 0 |
| **Nervous system disorders** | 1 (16.7) | 1 (16.7) | 0 | 1 (16.7) | 3 (12.5) | 1 (12.5) |
| Dizziness | 1 (16.7) | 0 | 0 | 0 | 1 (4.2) | 0 |
| Headache | 0 | 0 | 0 | 1 (16.7) | 1 (4.2) | 1 (12.5) |
| Muscle contractions involuntary | 0 | 1 | 0 | 0 | 1 (4.2) | 0 |
| **General disorders and administration site conditions** | 0 | 0 | 1 (16.7) | 0 | 1 (4.2) | 0 |
| Fatigue | 0 | 0 | 1 (16.7) | 0 | 1 (4.2) | 0 |
| **Infections and infestations** | 0 | 1 (16.7) | 0 | 0 | 1 (4.2) | 0 |
| Urinary tract infection | 0 | 1 (16.7) | 0 | 0 | 1 (4.2) | 0 |
| **Injury, poisoning and procedural complications** | 1 (16.7) | 0 | 0 | 0 | 1 (4.2) | 0 |
| Muscle strain | 1 (16.7) | 0 | 0 | 0 | 1 (4.2) | 0 |
| **Musculoskeletal and connective tissue disorders** | 0 | 0 | 1 (16.7) | 0 | 1 (4.2) | 0 |
| Arthralgia | 0 | 0 | 1 (16.7) | 0 | 1 (4.2) | 0 |

## Pharmacokinetics

### Plasma Concentrations of rhu-pGSN

Among the 32 subjects who were randomized with rhu-pGSN (N=24) or placebo (N=8), one subject in the 18 mg/kg group had a major protocol deviation that led to exclusion of her data from the Per Protocol (PP) population. Summary statistics of pGSN plasma concentration and the PK parameters are presented only for subjects who were part of both the PK and PP populations.

The mean baseline pGSN plasma concentrations (pre-treatment on Day 1, representing endogenous pGSN) for the 6, 12, 18, and 24 mg/kg cohorts were: 64.8, 63.6, 59.6, and 72.5 µg/mL, respectively. The mean pGSN plasma concentration for the placebo cohort at baseline was 70.0 µg/mL (Supplementary Table 3). For each individual, concentrations above the baseline endogenous pGSN level were calculated by subtracting the pre-dose pGSN level from each subsequent observed concentration. Baseline-adjusted concentrations that were < 0 were set to zero. Mean baseline-adjusted plasma rhu-pGSN concentrations were illustrated by dose group in Fig. 3A-C. The mean maximum plasma rhu-pGSN concentrations were observed within 15 minutes after completion of treatment with the exception of Cohort 3 (18 mg/kg) which reached mean maximum plasma rhu-pGSN concentrations after Doses 1 and 2 at 1 hour after completion of treatment. Mean maximum plasma rhu-pGSN concentrations declined over time across cohorts, except for Cohort 3 following Dose 1 and 2, in which the 4-hour post-dose concentrations exceeded those observed at 2 hours. For all other cohorts and dose groups, plasma concentrations decreased gradually at each time point over the 12- or 24-hours post-treatment. The mean 24-hour post-dose rhu-pGSN plasma concentrations following Dose 2 and Dose 5 are summarized in Table 4. An overall dose-dependent increase in 24-hour post-dose concentrations was observed, except between the 18 mg/kg and 24 mg/kg cohorts after Dose 2, which may be due to high variability in the 18 mg/kg cohort after Dose 2 (Supplementary Table 5). Mean plasma rhu-pGSN concentrations in all cohorts had returned to near-baseline levels by Day 28 (Table 4).

**Fig. 3.**
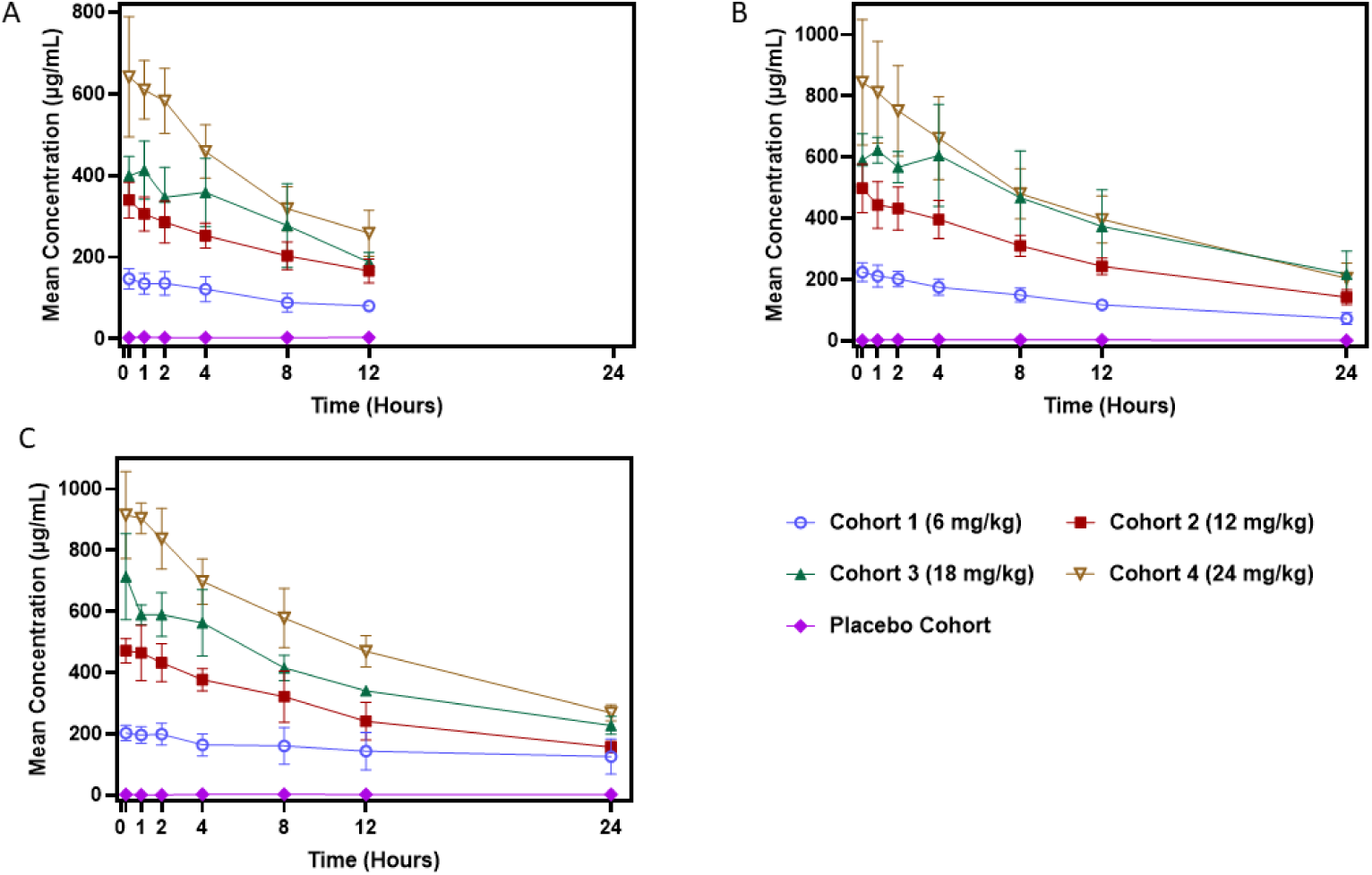
Mean baseline-adjusted plasma rhu-pGSN concentrations following dose 1 (A), dose 2 (B) and dose 5 (C) in subjects who were part of both the PK and PP populations

**Table 4.** Mean baseline-adjusted plasma concentrations of rhu-pGSN at 24 hours post-dose, and on Day 8 and 28 in subjects who were part of both the PK and PP populations.

| Time Points | rhu-pGSN |  |  |  | Placebo<br>N=8 |
| --- | --- | --- | --- | --- | --- |
|  | 6 mg/kg<br>N=6 | 12 mg/kg<br>N=6 | 18 mg/kg<br>N=5<sup>a</sup> | 24 mg/kg<br>N=6 |  |
| 24-hour post Dose 2 | 72.3 (19.3) | 142.7 (24.5) | 218.0 (74.6) | 204.5 (50.1) | 1.6 (4.6) |
| 24-hour post Dose 5 | 125.9 (58.0) | 156.9 (19.4) | 228.0 (29.5) | 269.4 (26.2) | 2.7 (5.4) |
| Day 8 | 36.0 (23.0) | 40.5 (16.2) | 46.4 (23.6) | 46.9 (24.2) | 6.1 (6.7) |
| Day 28 | 19.7 (16.3) | 8.4 (9.5) | 18.5 (7.7) | 9.3 (10.7) | 4.2 (6.1) |

<sup>a</sup> One subject in the 18 mg/kg group had a major protocol deviation that led to exclusion of her data from the Per Protocol (PP) population.

### Pharmacokinetic Parameters of rhu-pGSN

Key PK parameters of baseline-adjusted rhu-pGSN were summarized in Table 5. Mean PK parameters of rhu-pGSN showed higher exposure (AUC_0-8_, AUC_0-12_, AUC_0-24_, and AUC_0-inf_) with increasing dose except for AUC_0-inf_ in Cohort 3 (18 mg/kg) after Dose 2 which was slightly higher than the exposure observed in Cohort 4 (24 mg/kg) (13947.5 h* μg/mL and 13682.4 h* μg/mL, respectively). This may be due to high variability in the 18 mg/kg cohort after Dose 2 (Supplementary Table 5). Mean t_1/2_ (SD) varied from 9.2 (3.0) to 28.4 (8.5) hours across all doses and cohorts and the median t_1/2_ was 14.5 h in all dosing regimens, but sampling was limited to 24-h postdose measurements. The rhu-pGSN level rose well above baseline at C_max_ for each dose, with the higher doses (18 mg/kg and 24 mg/kg) yielding pGSN levels more than 10 times the baseline values.

**Table 5.** Summary of mean pharmacokinetics parameters in subjects who were part of both the PK and PP populations.

| Geometric Mean<br>Parameters (SD) | rhu-pGSN |  |  |  |
| --- | --- | --- | --- | --- |
|  | 6 mg/kg<br>N=6 | 12 mg/kg<br>N=6 | 18 mg/kg<br>N=5<sup>a</sup> | 24 mg/kg<br>N=6 |
| **Dose 1** |  |  |  |  |
| C<sub>max</sub> (µg/mL)<sup>b</sup> | 148.4 (26.8) | 347.5 (41.2) | 442.5 (50.2) | 663.6 (110.8) |
| AUC<sub>0-8</sub> (h*µg/mL)<sup>b</sup> | 912.4 (201.9) | 2010.0 (261.8) | 2659.9 (474.3) | 2695.6 (475.1) |
| AUC<sub>0-12</sub> (h*µg/mL)<sup>b</sup> | 1245.5 (257.9) | 2737.5 (379.1) | 3547.0 (657.3) | 4832.3 (646.6) |
| t<sub>1/2</sub> (h) | 14.0 (5.4) | 13.1 (2.6) | 9.2 (3.0) | 10.7 (5.2) |
| **Dose 2** |  |  |  |  |
| C<sub>max</sub> (µg/mL)<sup>b</sup> | 225.2 (27.0) | 506.9 (78.5) | 691.5 (110.5) | 855.7 (200.6) |
| AUC<sub>0-8</sub> (h*µg/mL)<sup>b</sup> | 1422.9 (179.6) | 3089.9 (388.7) | 4389.6 (851.8) | 5132.3 (1013.6) |
| AUC<sub>0-12</sub> (h*µg/mL)<sup>b</sup> | 1953.6 (234.5) | 4194.6 (448.6) | 6022.1 (1376.2) | 6859.8 (1318.6) |
| AUC<sub>0-24</sub> (h*µg/mL)<sup>b</sup> | 3066.2 (256.8) | 6448.5 (599.5) | 9374.2 (2456.0) | 10258.2 (2035.6) |
| AUC<sub>0-inf</sub> (h*µg/mL)<sup>b</sup> | 4857.8 (933.3) | 9394.8 (1641.5) | 13947.5 (4245.5) | 13682.4 (3094.3) |
| t<sub>1/2</sub> (h) | 16.7 (6.1) | 14.5 (3.4) | 15.1 (4.5) | 12.0 (1.5) |
| **Dose 5** |  |  |  |  |
| C<sub>max</sub> (µg/mL)<sup>b</sup> | 218.9 (37.7) | 501.6 (60.4) | 703.8 (140.7) | 970.9 (87.9) |
| AUC<sub>0-8</sub> (h*µg/mL)<sup>b</sup> | 1371.8 (252.5) | 3033.3 (329.1) | 4257.9 (561.1) | 5750.0 (539.1) |
| AUC<sub>0-12</sub> (h*µg/mL)<sup>b</sup> | 1958.7 (463.3) | 4131.6 (579.1) | 5770.0 (632.3) | 7824.6 (816.2) |
| AUC<sub>0-24</sub> (h*µg/mL)<sup>b</sup> | 3486.5 (1035.2) | 6599.1 (1334.2) | 9139.1 (793.1) | 12135.3 (1172.4) |
| AUC<sub>0-inf</sub> (h*µg/mL)<sup>b</sup> | 6634.8 (2055.4) | 12006.2 (7502.8) | 14909.2 (1294.6) | 17779.1 (1810.9) |
| t<sub>1/2</sub> (h) | 28.4 (8.5) | 22.0 (11.3) | 17.6 (1.9) | 14.5 (2.2) |

<sup>a</sup> One subject in the 18 mg/kg group had a major protocol deviation that led to exclusion of her data from the Per Protocol (PP) population.
<sup>b</sup> Baseline pGSN concentrations (pre-Dose 1, Day 1) were subtracted from each observed concentration of each subject.

### Immunogenicity

At day 1 (predose), 4 out of 32 subjects were screened positive in the Tier I, but none of them was confirmed positive for the presence of anti-pGSN antibodies in the Tier II (Table 6). On Day 28, 18 subjects were screened positive for the presence of anti-pGSN antibodies in Tier I, 2 additional samples had replicate results which spanned the cut-off point. Among the 20 samples requiring Tier II analysis, 18 were confirmed negative, 1 in the placebo group was confirmed positive, 1 in the 24 mg/kg rhu-pGSN group has a non-reportable result. The two samples were further tested in Tier III, but both show near-baseline signals at the lowest dilution tested (30-fold). Therefore, it is reasonable to consider those two samples were antibody negative.

**Table 6.**
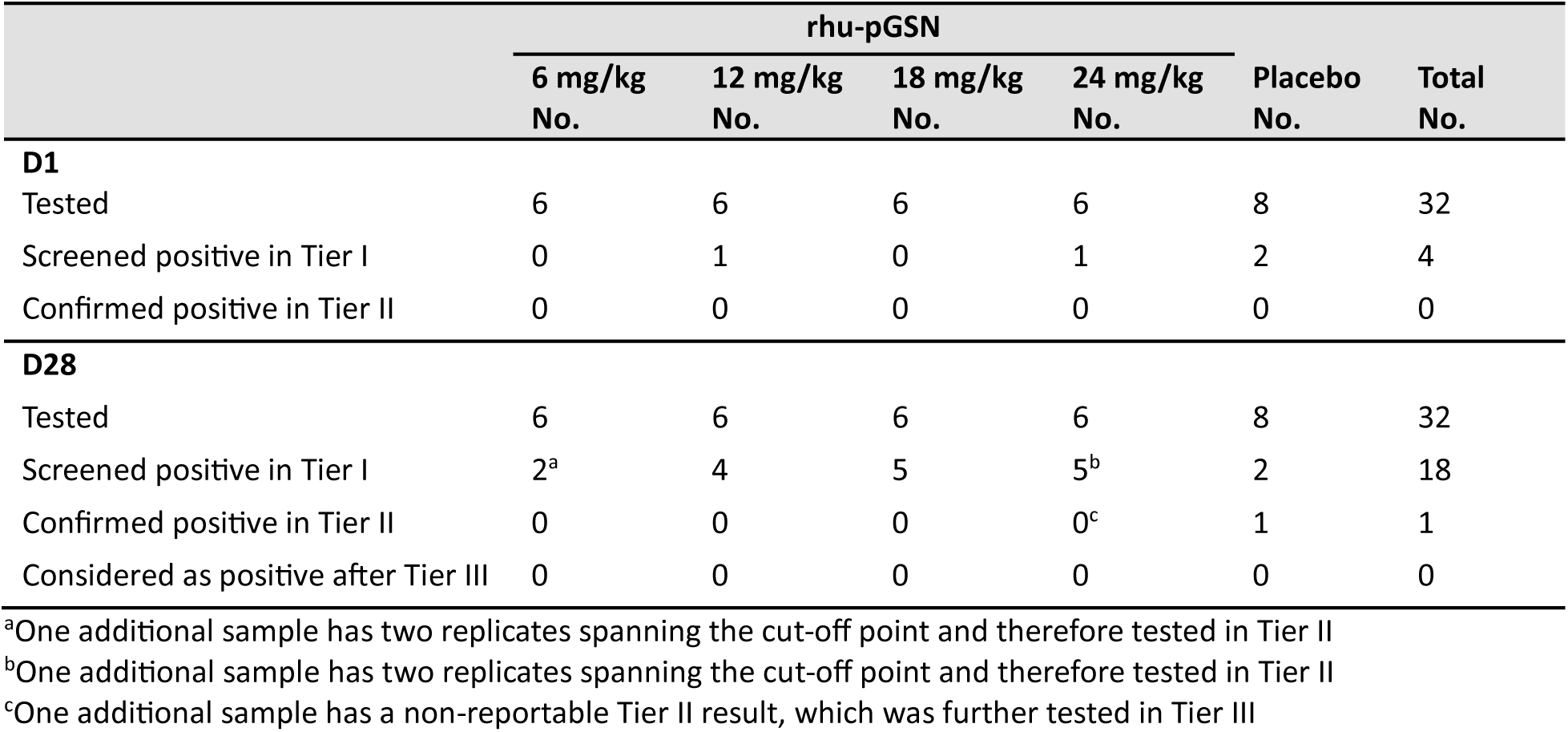
Frequencies of anti-drug antibodies at day 1 (predose) and day 28 in different cohorts.

|  | rhu-pGSN |  |  |  | Placebo<br>No. | Total<br>No. |
| --- | --- | --- | --- | --- | --- | --- |
|  | 6 mg/kg<br>No. | 12 mg/kg<br>No. | 18 mg/kg<br>No. | 24 mg/kg<br>No. |  |  |
| **D1** |  |  |  |  |  |  |
| Tested | 6 | 6 | 6 | 6 | 8 | 32 |
| Screened positive in Tier I | 0 | 1 | 0 | 1 | 2 | 4 |
| Confirmed positive in Tier II | 0 | 0 | 0 | 0 | 0 | 0 |
| **D28** |  |  |  |  |  |  |
| Tested | 6 | 6 | 6 | 6 | 8 | 32 |
| Screened positive in Tier I | 2<sup>a</sup> | 4 | 5 | 5<sup>b</sup> | 2 | 18 |
| Confirmed positive in Tier II | 0 | 0 | 0 | 0<sup>c</sup> | 1 | 1 |
| Considered as positive after Tier III | 0 | 0 | 0 | 0 | 0 | 0 |

<sup>a</sup>One additional sample has two replicates spanning the cut-off point and therefore tested in Tier II
<sup>b</sup>One additional sample has two replicates spanning the cut-off point and therefore tested in Tier II
<sup>c</sup>One additional sample has a non-reportable Tier II result, which was further tested in Tier III

## Discussion

We assessed the safety and tolerability of rhu-pGSN in a phase 1, randomized, double-blind, placebo-controlled, multiple-dose escalation study. Prior to this study, IV administration of rhu-pGSN had been evaluated by BioAegis Therapeutics in two clinical trials in hospitalized patients: one in patients with mild community-acquired pneumonia (CAP) and another in patients with moderate to severe SARS-CoV-2(COVID-19) (27,28). In the CAP trial, 33 subjects admitted to hospital with mild CAP received 1 to 3 doses of 6 −24 mg/kg of rhu-pGSN or placebo over 3 days. All tested doses of rhu-pGSN were generally well tolerated in this study, with no drug-related AEs in the rhu-pGSN arms. Two subjects died during the study: one received a single dose of the lowest dose of rhu-pGSN, the other received placebo (27). The COVID-19 trial enrolled 61 patients with World Health Organization (WHO) severity score of 4-6 to receive adjunctive rhu-pGSN or placebo on top of standard-of-care. Subjects in the rhu-pGSN arm received three doses of 12 mg/kg rhu-pGSN at entry, 12 hours and 36 hours. Rhu-pGSN was well tolerated, with numerically fewer subjects experiencing SAEs in the rhu-pGSN group (5 subjects; 16.7%) than in the placebo group (8 subjects; 25.8%) (28).

The current study significantly expanded the safety profile characterization of pGSN beyond that achieved in previous studies which were limited to up to three doses. In the current study, the safety, tolerability, and pharmacokinetics of five doses of rhu-pGSN, ranging from 6 mg/kg to 24 mg/kg, were evaluated in healthy volunteers. Doses of rhu-pGSN or placebo were IV administered at 0 h, 12 h, 36 h, 60 h and 84 h. Infusion with increasing doses of rhu-pGSN yielded a sustained dose-dependent increase in pGSN levels in the circulation. Repeated dosing did not lead to significant drug accumulation. In the higher rhu-pGSN dosing cohorts (18 and 24 mg/kg), peak pGSN concentrations exceeded 10 times the physiological level, without any SAEs or AEs that led to study discontinuation or interruption. The half-life of rhu-pGSN in different dosing regimens varied between 9.2 h and 28.4 h, with the median half-life of rhu-pGSN exceeding 14 h across all dosing regimens, supporting once daily IV dosing in the healthy subjects studied in this trial. A total of 13 AEs were reported by 10 of 24 subjects (41.7%) who received rhu-pGSN, whereas 1 AE was reported by 1 of 8 subject (12.5%) who received placebo. All AEs were mild or moderate. No apparent association was observed between the rhu-pGSN dose and the frequency or types of AEs reported.

## Conclusions

Overall, IV administration of five doses of rhu-pGSN up to 24 mg/kg over 5 days was safe and well tolerated in this small trial in healthy volunteers. In the higher dosing cohorts, even with pGSN levels maintained well above physiological concentration over the 5 days, neither SAEs nor AEs that led to study discontinuation/interruption were observed. The median half-life of rhu-pGSN exceeded 14 h across all dosing regimens, supporting once daily IV dosing in healthy subjects. A phase 2 proof-of-concept trial evaluating the efficacy and safety of rhu-pGSN in addition to standard of care in subject with moderate-to severe ARDS due to pneumonia or other infections is currently ongoing.

## Supporting information

Supplemental file 1 - Study Protocol revised

Supplemental file 2 - Inclusion and Exclusion Criteria and Supplementary Table 1-6

## Data Availability

All data produced in the present study are available upon reasonable request to the authors

## List of abbreviations

AE: adverse events
ALI: acute lung injury
ARDS: acute respiratory distress syndrome
BMI: body mass index
CAP: community-acquired pneumonia
EKG: electrocardiogram
ELISA: enzyme-linked immunosorbent assay
EOS: End of Study
IRB: Institutional Review Board
IV: intravenous
MedDRA: Medical Dictionary for Regulatory Activities
PK: pharmacokinetic(s)
PP: Per Protocol
rhu-pGSN: recombinant human plasma gelsolin
SAE: serious adverse event
TEAE: treatment-emergent adverse event

## Declarations

## Human ethics approval and consent to participate declaration

The study was designed and conducted in accordance with the Declaration of Helsinki and was approved by Advarra’ s Institutional Review Board (IRB# Pro00069641) before the start of the trial. Written informed consent was obtained from all participants.

## Consent for publication

Not applicable

## Availability of data and materials

The datasets used and/or analyzed during the current study are available from the corresponding author on reasonable request.

## Authors’ contribution

YL analyzed and interpreted the data, and wrote the first draft of the manuscript. SL and MJD contributed to the design, execution and data analysis of the clinical trial. EK and JP performed the plasma gelsolin ELISA. LK contributed to manuscript writing. All coauthors reviewed and approved the final version of the manuscript.

## Acknowledgements

Recombinant human plasma gelsolin was provided by BioAegis Therapeutics.

We thank Nucleus Network for invaluable help conducting the trial and Jennifer Stanley for CSR preparation.

We also thank all the healthy volunteers who participated in the trial, as well as the principal investigator at the study site: Trisha Shamp (Saint Paul, Minnesota).

## References

1. Smith DB, Janmey PA, Herbert TJ, Lind SE. Quantitative measurement of plasma gelsolin and its incorporation into fibrin clots. J Lab Clin Med. 1987 Aug;110(2):189–95.

2. Ordija CM, Chiou TTY, Yang Z, Deloid GM, de Oliveira Valdo M, Wang Z, et al. Free actin impairs macrophage bacterial defenses via scavenger receptor MARCO interaction with reversal by plasma gelsolin. American Journal of Physiology–Lung Cellular and Molecular Physiology. 2017;312(6):L1018–28.

3. Yang Z, Chiou TTY, Stossel TP, Kobzik L. Plasma gelsolin improves lung host defense against pneumonia by enhancing macrophage NOS3 function. American Journal of Physiology–Lung Cellular and Molecular Physiology. 2015;309(1):L11–6.

4. Osborn TM, Dahlgren C, Hartwig JH, Stossel TP. Modifications of cellular responses to lysophosphatidic acid and platelet-activating factor by plasma gelsolin. American Journal of Physiology-Cell Physiology. 2007;292(4):C1323–30.

5. Bucki R, Georges PC, Espinassous Q, Funaki M, Pastore JJ, Chaby R, et al. Inactivation of endotoxin by human plasma gelsolin. Biochemistry. 2005;44(28):9590–7.

6. Lind SE, Smith DB, Janmey PA, Stossel TP. Role of plasma gelsolin and the vitamin D-binding protein in clearing actin from the circulation. The Journal of Clinical Investigation. 1986;78(3):736–42.

7. Bhopale VM, Ruhela D, Brett KD, Nugent NZ, Fraser NK, Levinson SL, et al. Plasma gelsolin modulates the production and fate of IL-1β-containing microparticles following high-pressure exposure and decompression. Journal of Applied Physiology. 2021;130(5):1604–13.

8. Sharma RK, Goswami B, Das Mandal S, Guha A, Willard B, Ray PS. Quorum sensing by gelsolin regulates programmed cell death 4 expression and a density-dependent phenotype in macrophages. The Journal of Immunology. 2021;207(5):1250–64.

9. Self WH, Wunderink RG, DiNubile MJ, Stossel TP, Levinson SL, Williams DJ, et al. Low admission plasma gelsolin concentrations identify community-acquired pneumonia patients at high risk for severe outcomes. Clinical Infectious Diseases. 2019;69(7):1218–25.

10. Mounzer KC, Moncure M, Smith YR, DiNubile MJ. Relationship of admission plasma gelsolin levels to clinical outcomes in patients after major trauma. American Journal of Respiratory and Critical Care Medicine. 1999;160(5):1673–81.

11. Güntert A, Campbell J, Saleem M, O’ Brien DP, Thompson AJ, Byers HL, et al. Plasma gelsolin is decreased and correlates with rate of decline in Alzheimer’ s disease. Journal of Alzheimer’ s Disease. 2010;21(2):585–96.

12. Kulakowska A, Drozdowski W, Sadzynski A, Bucki R, Janmey PA. Gelsolin concentration in cerebrospinal fluid from patients with multiple sclerosis and other neurological disorders. European Journal of Neurology. 2008;15(6):584–8.

13. Osborn TM, Verdrengh M, Stossel TP, Tarkowski A, Bokarewa M. Decreased levels of the gelsolin plasma isoform in patients with rheumatoid arthritis. Arthritis Research & Therapy. 2008;10(5):R117.

14. Abers MS, Delmonte OM, Ricotta EE, Fintzi J, Fink DL, Jesus AAA de, et al. An immune-based biomarker signature is associated with mortality in COVID-19 patients. JCI Insight [Internet]. 2021 Jan 11 [cited 2026 Jan 23];6(1). Available from: https://insight.jci.org/articles/view/144455

15. Messner CB, Demichev V, Wendisch D, Michalick L, White M, Freiwald A, et al. Ultra-high-throughput clinical proteomics reveals classifiers of COVID-19 infection. Cell Systems. 2020;11(1):11–24.

16. Overmyer KA, Shishkova E, Miller IJ, Balnis J, Bernstein MN, Peters-Clarke TM, et al. Large-scale multi-omic analysis of COVID-19 severity. Cell Systems. 2021;12(1):23–40.

17. Shi S, Chen C, Zhao D, Liu X, Cheng B, Wu S, et al. The role of plasma gelsolin in cardiopulmonary bypass–induced acute lung injury in infants and young children: a pilot study. BMC Anesthesiology. 2014;14(1):67.

18. DiNubile MJ, Stossel TP, Ljunghusen OC, Ferrara JL, Antin JH. Prognostic implications of declining plasma gelsolin levels after allogeneic stem cell transplantation. Blood. 2002;100(13):4367–71.

19. Holm FS, Sivapalan P, Seersholm N, Itenov TS, Christensen PH, Jensen JUS. Acute lung injury in critically ill patients: actin-scavenger gelsolin signals prolonged respiratory failure. Shock. 2019;52(3):370–7.

20. Lee PS, Waxman AB, Cotich KL, Chung SW, Perrella MA, Stossel TP. Plasma gelsolin is a marker and therapeutic agent in animal sepsis. Critical Care Medicine. 2007;35(3):849–55.

21. Lee J, Sasaki F, Koike E, Cho M, Lee Y, Dho SH, et al. Gelsolin alleviates rheumatoid arthritis by negatively regulating NLRP3 inflammasome activation. Cell Death & Differentiation. 2024;31(12):1679–94.

22. Bhat AR, Arya AK, Bhopale VM, Imtiyaz Z, Thom SR. Recombinant human plasma gelsolin suppresses persistent neuroinflammation and restores hippocampal neurogenesis in murine model of decompression sickness. Journal of Neurophysiology. 2024;132(6):1877–86.

23. Arya AK, Sethuraman K, Waddell J, Cha YS, Liang Y, Bhopale VM, et al. Inflammatory responses to acute carbon monoxide poisoning and the role of plasma gelsolin. Sci Adv. 2025 Feb 7;11(6):eado9751.

24. DiNubile MJ, Levinson SL, Stossel TP, Lawrenz MB, Warawa JM. Recombinant human plasma gelsolin improves survival and attenuates lung injury in a murine model of multidrug-resistant Pseudomonas aeruginosa pneumonia. Open Forum Infectious Diseases. 2020;7(8):ofaa236.

25. Yang Z, Bedugnis A, Levinson S, DiNubile M, Stossel T, Lu Q, et al. Delayed administration of recombinant plasma gelsolin improves survival in a murine model of penicillin-susceptible and penicillin-resistant pneumococcal pneumonia. The Journal of Infectious Diseases. 2019;220(9):1498–502.

26. Cui TX, Brady AE, Zhang YJ, Fulton CT, Popova AP. Gelsolin attenuates neonatal hyperoxia-induced inflammatory responses to rhinovirus infection and preserves alveolarization. Frontiers in Immunology. 2022;13:792716.

27. Tannous A, Levinson SL, Bolognese J, Opal SM, DiNubile MJ. Safety and Pharmacokinetics of Recombinant Human Plasma Gelsolin in Patients Hospitalized for Nonsevere Community-Acquired Pneumonia. Antimicrobial Agents and Chemotherapy. 2020 Sept 21;64(10):10.1128/aac.00579-20.

28. DiNubile MJ, Parra S, Salomó AC, Levinson SL. Adjunctive Recombinant Human Plasma Gelsolin for Severe Coronavirus Disease 2019 Pneumonia. Open Forum Infectious Diseases. 2022 Aug 2;9(8):ofac357.

