## Supplemental file 1 - Study Protocol revised for "A Phase 1, Single-Center, Randomized, Double-Blind, Placebo-Controlled, Multiple-Dose Escalation Study for the Evaluation of the Safety, Tolerability, and Pharmacokinetics of Recombinant Human Plasma Gelsolin (rhu-pGSN) Following Intravenous Administration to Healthy Volunteers"

| | |
|---|---|
| **Protocol Number:** | BTI-101 |
| **Investigational Product:** | rhu-pGSN |
| **IND Number:** | 163130 |
| **EUDRA CT Number** | N/A |
| **Development Phase:** | Phase 1 |
| **Indication Studied:** | Healthy volunteers |
| **Sponsor Name and Address:** | BioAegis Therapeutics, Inc. (BTI) |
| **Responsible Medical Officer:** | Mark J. DiNubile, MD FIDSA |
| **Compliance Statement:** | This study will be conducted in accordance with the ethical principles that have their origin in the Declaration of Helsinki, clinical research guidelines established by the Code of Federal Regulations (Title 21, CFR Parts 50, 56, and 312), and ICH GCP Guidelines. Essential study documents will be archived in accordance with applicable regulations. |
| **Protocol Date:** | 31 January 2023 |
| **Version:** | 1.2 |

### PROTOCOL APPROVAL SIGNATURE PAGE

| | |
|---|---|
| **Protocol:** | BTI-101 |
| **Title:** | A phase 1, single-center, randomized, double-blind, placebo-controlled, multiple ascending dose escalation, parallel design study for the evaluation of the safety, tolerability, and pharmacokinetics of recombinant human plasma gelsolin (rhu-pGSN) following intravenous administration to healthy volunteers |
| **Date:** | 31 January 2023 |
| **Amendment:** | N/A |

Reviewed and Approved by:

| | |
|---|---|
| Mark J. DiNubile, MD<br>Chief Medical Officer<br>BioAegis Therapeutics, Inc. | January 31, 2023<br>Date |

### PROTOCOL ACCEPTANCE FORM

| | |
|---|---|
| **Protocol:** | BTI-101 |
| **Title:** | A phase 1, single-center, randomized, double-blind, placebo-controlled, multiple ascending dose escalation, parallel design study for the evaluation of the safety, tolerability, and pharmacokinetics of recombinant human plasma gelsolin (rhu-pGSN) following intravenous administration to healthy volunteers |
| **Date:** | 31 January 2023 |
| **Amendment:** | N/A |

I have carefully read the BTI-101 protocol and agree that it contains all of the necessary information required to conduct this study. I agree to conduct this study as described and according to the Declaration of Helsinki, ICH Guidelines for GCP, and all applicable regulatory requirements.

________________________________________ ________________

Investigator’s Signature Date

________________________________________

Name (printed)

### 1. SYNOPSIS

| | |
|---|---|
| **Name of Sponsor/Company:**<br>BioAegis Therapeutics Inc. | |
| **Name of Investigational Product:**<br>Recombinant Human Plasma Gelsolin (rhu-pGSN) | |
| **Name of Active Ingredient:**<br>rhu-pGSN | |
| **Title of Study:**<br>A phase 1, single-center, randomized, double-blind, placebo-controlled, multiple ascending dose escalation, parallel design study for the evaluation of the safety, tolerability, and pharmacokinetics of recombinant human plasma gelsolin (rhu-pGSN) following intravenous administration to healthy volunteers | |
| **Study Center(s):** 1 site | |
| **Studied Period (Years):**<br>Estimated date first subject enrolled: MARCH 2023<br>Estimated date last subject completed: MAY 2023 | **Phase of development:**<br>1 |
| **Objectives:**<br>**Primary**<br>• To evaluate the safety and tolerability of 4 sequentially ascending doses of rhu-pGSN at 6 mg/kg, 12 mg/kg, 18 mg/kg, and 24 mg/kg of actual body weight administered by intravenous (IV) infusion for 5 doses at 0, 12, 36, 60, and 84 hours to healthy subjects<br>**Secondary**<br>• To characterize the pharmacokinetic (PK) profile of rhu-pGSN following administration of multiple (5) doses of rhu-pGSN to healthy subjects assigned to 4 dose levels (6, 12, 18, or 24 mg/kg) [PK specimens are to be obtained for doses 1, 2, and 5; post-dose samples are timed from the end of the infusions]<br>• To investigate the development of anti-pGSN antibodies following administration of multiple (5) doses of rhu-pGSN administered to healthy subjects assigned to 4 dose levels (6, 12, 18, or 24 mg/kg) | |
| **Study Design:**<br>Study BTI-101 is a Phase 1, randomized, double-blind, placebo-controlled, dose escalation, parallel design study to evaluate the safety, tolerability, and PK of IV rhu-pGSN or saline placebo administered as 5 doses each of 6, 12, 18, or 24 mg/kg of body weight. Each of 4 dosing cohorts will include 8 subjects randomized 3:1 rhu-pGSN:placebo (6 rhu-pGSN subjects:2 placebo subjects). Subjects will be healthy adults 18-50 years of age.<br>Doses will be administered at 0 hours (Day 1), 12 hours (Day 1), 36 hours (Day 2), 60 hours (Day 3), and 84 hours (Day 4). Subjects will be kept in-house until after the last blood sample is taken on Day 5. Subjects will return for follow-up 7 days after the initiation of therapy (Day 8) and on Day 28 for the End-of-Study (EOS) Visit. After each cohort has completed the Day 8 visit, review of the safety results (including labs) will be conducted (and unblinded where appropriate) before the initiation of the next higher dose cohort.<br>To assess safety and tolerability, subjects will undergo physical examinations (including vital sign measurements), adverse event (AE) assessments, concomitant medication assessments, and safety | |

laboratory testing. Blood samples will be collected for analysis of pGSN levels and antibodies against pGSN.

**Number of Subjects (Enrolled):**

A total of 32 (6 rhu-pGSN and 2 placebo recipients per dose cohort × 4 dosing levels)

**Inclusion Criteria:**

1. Healthy male or female adults 18 to 55 years of age without chronic or active acute medical conditions
2. Informed consent obtained from subject
3. Weight ≤100 kg and body mass index (BMI) <30 kg/m$^2$
4. Willingness during the course of the study, starting at screening and for at least 3 months after their final study treatment:
   a) Female subjects of childbearing potential must agree to use 2 medically accepted/Food and Drug Association (FDA)-approved birth control methods
   b) Male subjects with a partner who might become pregnant must agree to use reliable forms of contraception (i.e., condom, vasectomy, abstinence), and an acceptable method of birth control must be used by the partner
   c) All subjects must agree not to donate sperm or eggs

**Exclusion Criteria:**

1. Pregnant or lactating women
2. Acute illness during the month prior to screening
3. Circumstances that may require any medications (including prescription medication, over-the-counter medication, vitamins, or supplements) during the 5 inpatient days of the study other than acetaminophen
4. Hospitalization during the year prior to screening
5. History of cancer or treatment with systemic chemotherapy or radiation therapy at any time
6. Transplantation of hematopoietic or solid organs
7. History of diabetes mellitus; myocardial infarction, angina, or other cardiovascular disease; stroke or cerebrovascular disease; chronic obstructive pulmonary disease (COPD) or asthma; deep vein thrombosis (DVT)/pulmonary embolism (PE); liver or kidney disease; psychiatric condition; or active or chronic infection
8. Receipt of blood products during the year prior to screening
9. Chronic mechanical ventilation or dialysis
10. Any clinically significant abnormalities in clinical chemistry, hematology, or urinalysis results as judged by the Investigator
11. Any clinically significant abnormalities of vital signs, EKG or physical examination findings as judged by the Investigator
12. Positive results for recreational drugs during screening
13. Any other condition deemed by the Investigator as possibly interfering with the conduct of the study

**Investigational Product, Dosage, and Mode of Administration:**

rhu-pGSN dissolved in sterile water infused IV at a rate of ≤20 mL per minute through a standard 0.20 µ filter at a dose of 6 mg/kg, 12 mg/kg, 18 mg/kg, or 24 mg/kg. Each dose level is administered at 5 time points: 0, 12 (Day 1), 36 (Day 2), 60 (Day 3), and 84 hours (Day 4).

**Duration of Treatment:**

5 doses over 4 days (84 hours)

**Duration of Subject Participation:**

Subject screening: up to 6 weeks. Study treatment: 4 days. Safety follow-up: through 28 days post-first dose. Maximum duration: 10 weeks.

**Reference Therapy, Dosage, and Mode of Administration:**

Placebo will be dosed at a weight-based volume of 0.025 mL/mg × dose in mg/kg × weight in kg of 0.9% saline solution via IV push (matching the volume of rhu-pGSN for a subject of that size) injected at ≤20 mL/minute though a standard 0.2 μ filter. Placebo will be administered on the same schedule as the active drug.

**Endpoints/Outcomes:**

**Primary**

- Incidence and severity of clinical and laboratory AEs regardless of causality graded according to the *Toxicity Grading Scale for Healthy Adult and Adolescent Volunteers Enrolled in Preventive Vaccine Clinical Trials*

**Secondary**

- PK for IV rhu-pGSN including, but not limited to: $C_{max}$, $T_{max}$, $T_{1/2}$, $AUC_{0-8}$, and additionally after Doses #2 and #5, $AUC_{0-24}$, and $AUC_{inf}$ (levels measured on Day 1 within 30 minutes predose, and at 15 minutes and 1, 2, 4, 8, and 12 hours after Dose #1 and on Days 1 and 2 and on Days 4 and 5 within 30 minutes predose, and at 15 minutes and 1, 2, 4, 8, 12, and 24 hours after Doses #2 and #5) (post-dose samples are timed from the end of the infusion]
- Presence of anti-rhu-pGSN antibodies at Day 28

**Safety Review:**

The safety data will be reviewed by a Safety Review Committee (SRC) comprised of at least 1 expert clinician, the Medical Monitor at the contract research organization, and the Sponsor's Chief Medical Officer. All AEs occurring during this healthy-volunteer study will be presumed to be related to study drug unless occurring before the study therapy is initiated pending review by the SRC.

Adverse events (AEs) will be categorized as "serious" according to the judgement of the site investigator and then reviewed and adjudicated by the SRC. The "severity" of an AE will be graded according to the *Toxicity Grading Scale for Healthy Adult and Adolescent Volunteers Enrolled in Preventive Vaccine Clinical Trials*. An individual patient will be discontinued permanently from study treatment if they experience any serious adverse event (SAE) or a severity Grade 3 or greater AE (including infusion reactions). Further study treatment and enrollment will be paused pending review by the SRC if there are two Grade 3 AEs, one Grade 4 AE, or one SAE. The study will only be resumed if the SRC unanimously judges the AEs as unrelated to study treatment and procedures after consultation with regulatory and outside experts as needed.

The SRC will review safety data (including lab tests) through the Day 7 visit for each dosing arm to determine if it is safe to initiate the next cohort according to the safety criteria for stopping doses in the full protocol.

**Statistical Methods and Sample Size Rationale for Power Calculations:**

For each dosing group, 8 subjects will be randomized in a 3:1 ratio to receive an equal volume of rhu-pGSN or saline placebo during the treatment period. Baseline characteristics, safety, PK, and development of anti-drug antibodies (ADAs) results will be summarized via summary statistics separately for each rhu-pGSN dose level; data for all placebo subjects will be pooled. Additional details will be provided in a separate Statistical Analysis Plan (SAP).

Sample Size and Power Calculations

Prior studies of rhu-pGSN evaluated 3 doses of rhu-pGSN in subjects hospitalized with community-acquired pneumonia or coronavirus 2019 (COVID-19). To expand our understanding of possible rhu-pGSN toxicities in the absence of underlying disease, 24 healthy subjects will be administered 5 doses of rhu-pGSN at 6, 12, 18, and 24 mg/kg of body weight (6 subjects per dose group) and 8 healthy subjects (2 subjects per dose group) will be given the same volume on a weight basis of a visually indistinguishable 0.9% saline placebo to provide additional data to inform the safety and tolerability of rhu-pGSN.

The table below presents the minimum sample size such that there is a 90% probability of observing at least 1 AE of a certain type if the TRUE underlying AE incidence is as specified.

**Minimum Sample Size Calculations**

| Sample Size | TRUE underlying AE rate |
|---|---|
| 6 (each rhu-pGSN dose level) | 32% |
| 8 (pooled placebo) | 25% |
| 12 | 18% |
| 18 | 13% |
| 24 (pooled rhu-pGSN doses) | 10% |

With each sample size presented in the table above, if zero AEs of a certain type (e.g., serious adverse events [SAEs]) are observed, one could be "90% confident" that the TRUE underlying incidence for that AE is at most the percentage indicated above. Thus with 24 healthy subjects, if no SAEs are observed, the probability is 90% that the true underlying incidence of SAEs in healthy subjects administered 5 doses of rhu-pGSN at 6, 12, 18, or 24 mg/kg of body weight is at most 10%.

No data will be excluded from the summary statistics and analyses, and missing data will not be imputed. All subjects given at least part of 1 dose of study drug will be included in the intention-to-treat analysis (full analysis set). As a sensitivity analysis, a per-protocol analysis set excluding subjects who missed doses and/or randomly discontinued the study before the primary Day 7 Visit will be analyzed. The expectation is that all analysis sets will be almost identical.

Descriptive summaries of the incidence of all AEs, drug-related AEs, SAEs, drug-related SAEs, AEs leading to discontinuation, and deaths will be provided. All AEs and drug-related AEs will be summarized by counts and proportions of subjects having an AE, each AE type, and an AE of each System Organ Class. Serious AEs will be summarized similarly. AEs leading to discontinuation of treatment or the study will be summarized. All reasons for any early discontinuations will be summarized.

Laboratory data will be summarized in relation to normal range values via counts and percentages of subjects below, within, and above the respective normal range, and by summary statistics (N, mean, median, standard deviation, minimum, maximum, 90% confidence intervals, etc.) for baseline, each observed time point, and change from baseline at each observed time point for continuous lab endpoints.

Demographics and medical history characteristics will be summarized via counts and percentages of subjects for categorical variables, and by N, mean and standard deviation or median and interquartile range, as appropriate for the distribution of continuous variables.

$C_{max}$, $T_{max}$, $T_{1/2}$, $AUC_{0-8h}$, $AUC_{0-12}$, and additionally after Dose #5, $AUC_{0-24}$, and $AUC_{inf}$ will be calculated from the pGSN levels measured for Doses #1 and #5. Since the assay measures endogenous pGSN levels as well as exogenous recombinant pGSN, PK parameters will be calculated for values above baseline by subtracting predose pGSN level from each observed concentration. Tables and graphs will be provided.

Antibodies against rhu-pGSN will be assayed from frozen specimens to determine whether the investigational product induces an antibody response in recipients. The frequency of anti-pGSN antibodies with 95% confidence intervals will be provided for Day 1 (predose) and Day 28. If any subjects had antibodies at Day 1, the presence or absence of antibodies at Days 1 and 28 will be summarized in a 2 × 2 table (Day 1 vs. Day 28).

### 2. LIST OF ABBREVIATIONS AND DEFINITIONS OF TERMS

The following abbreviations and specialist terms are used in this study protocol.

**Table 1: Abbreviations and Specialist Terms**

| Abbreviation or Specialist Term | Explanation |
|---|---|
| ADA | anti-drug antibody |
| AE | adverse event |
| ALI | acute lung injury |
| ALT | alanine aminotransferase |
| aPTT | activated partial thromboplastin time |
| ARDS | acute respiratory distress syndrome |
| AST | aspartate aminotransferase |
| $AUC_{0-8}$ | area under the curve from time zero to 8 hours |
| $AUC_{inf}$ | area under the curve from time zero to infinity |
| BioAegis or BTI | BioAegis Therapeutics, Inc. or Sponsor |
| BMI | body mass index |
| CAP | community-acquired pneumonia |
| CBC | complete blood count |
| $C_{max}$ | maximum concentration |
| COPD | chronic obstructive pulmonary disease |
| COVID-19 | coronavirus 2019 |
| CPK | creatine phosphokinase |
| eCRF | electronic case report form |
| DSMB | Data and Safety Monitoring Board |
| DVT | deep vein thrombosis |
| EDC | electronic data capture |
| ELISA | enzyme-linked immunosorbent assay |
| EOS | end-of-study (visit) |
| f-actin | filamentous actin |
| FDA | Food and Drug Association |
| g-actin | globular actin |
| GCP | Good Clinical Practice |
| ICF | informed consent form |

| Abbreviation or Specialist Term | Explanation |
|---|---|
| ICH | International Council for Harmonization of Technical Requirements for Pharmaceuticals for Human Use |
| ICU | intensive care unit |
| IEC | Independent Ethics Committee |
| IRB | Institutional Review Board |
| IRR | infusion-related reaction |
| IUD | intrauterine device |
| IV | intravenous |
| MV | mechanical ventilation |
| NOAEL | no-observed-adverse-effect level |
| NSAIDs | non-steroidal anti-inflammatory drugs |
| NSS | normal saline solution |
| PDF | portable document format |
| PE | pulmonary embolism |
| PI | Principal Investigator<br>The investigator who leads the study conduct at an individual study site. Every study site has a principal investigator. |
| PK | pharmacokinetics |
| PT | prothrombin time |
| rhu-pGSN | recombinant human plasma gelsolin |
| SAE | serious adverse event |
| SAP | statistical analysis plan |
| SOC | standard of care |
| SRC | Safety Review Committee |
| SUSAR | suspected unexpected serious adverse reaction |
| $T_{1/2}$ | terminal half-life |
| TEAE | treatment-emergent adverse event |
| $T_{max}$ | time to maximum concentration |
| US | United States |
| VP | vasopressor |

### 3. INTRODUCTION

#### 3.1. Background

Acute hypoxemic respiratory failure can result from a severe inflammatory process in the lung (e.g., viral or bacterial pneumonia or inhalation injury) or distant from the lungs (e.g., intra-abdominal sepsis or necrotizing pancreatitis). In its advanced form (identified by a partial pressure of $O_2$/fraction of inspired $O_2$ [P/F ratio] <300, often <200), bilateral infiltrates develop that are not caused by volume overload or heart failure within 1 week of an inciting injury (often infection in the lung or a distant site), which culminates in intubation or noninvasive respiratory support. Using an expanded Berlin definition, these patients generally satisfy criteria for acute respiratory distress syndrome (ARDS) or at least moderate-to-severe acute lung injury (ALI) and require intensive care with mechanical or non-invasive ventilation or high-flow oxygen therapy. ALI/ARDS appears to be mediated by excessive and injurious inflammation that targets the lung whether or not the lung is or was the site of the initial injury.

#### 3.2. Current Treatment of ALI/ARDS

Other than supportive care and treatment of the precipitating event, no pharmacologic interventions aimed directly at the lung injury clearly benefit patients with ALI/ARDS. Interrupting or tempering the maladaptive inflammatory process could theoretically lessen further organ injury. To this end, glucocorticoids at various doses have been used without conclusive results in most circumstances. The evidence in ALI/ARDS associated with coronavirus 2019 (COVID-19) pneumonia indicates a role for steroids in severe-critical infections. Unfortunately, steroids induce an immunosuppressive state that may complicate ongoing infection or exaggerate the risk of nosocomial infection. Thus, there is an unmet need for less toxic alternative therapies for the treatment of inflammatory diseases leading to ALI/ARDS.

#### 3.3. Plasma Gelsolin

Plasma gelsolin is a human protein produced and secreted by virtually every cell type, and it circulates at high levels in the blood of healthy individuals. At normal levels of 200-300 mg/L, it is the fourth most abundant protein in the circulation.

The therapeutic protein being developed by BioAegis Therapeutics, Inc. (BioAegis or Sponsor) is recombinant human plasma gelsolin (rhu-pGSN). It is identical to the complete natural protein and is comprised of 755 amino acids. The protein, like the natural protein, is non-glycosylated. The protein consists of six repeated domains, which make up three distinct actin binding sites, two that bind globular actin (G-actin) and one that binds filamentous actin (F-actin). Domain 1 plus the first 10 amino acids of domain 2 of gelsolin is the minimal fragment size required for its actin severing activity. Rhu-pGSN contains multiple calcium binding sites that regulate its activity. It is a highly conserved protein, as far back as drosophila.

Plasma gelsolin modulates inflammation while at the same time, boosting the body’s ability to clear pathogens. It functions through mechanisms quite distinct from anti-inflammatory agents, which are antagonists of specific mediators or inhibitors of specific enzymes. Plasma gelsolin functions through a pleiotropic mechanism of action, scavenging toxic actin, binding inflammatory mediators and enhancing pathogen clearance. Repletion of pGSN in patients developing ALI/ARDS may limit lung injury and improve survival.

#### 3.4. Preclinical and Clinical Experience

##### 3.4.1. Preclinical Experience

Repeat IV administration of rhu-pGSN to rats at up to 25 or 100 mg/kg/day for 14 and 7 days, respectively, and to monkeys at up to 20 mg/kg/day for 14 days revealed no adverse effects on body weight, food consumption, clinical signs, ophthalmology parameters, clinical pathology, the cardiovascular system (monkeys), organ weights, and macroscopic and microscopic pathology. No clinically relevant pathological effects were evident in rats or monkeys following administration of rhu-pGSN for 14 days by IV injection. In addition, a study in Sprague Dawley rats utilizing the recently manufactured lot of BioAegis drug product administered at 100 mg/kg in 5 doses over 72 hours was conducted with no remarkable findings or adverse effects on the central nervous system. Thus, the no-observed-adverse-effect levels (NOAELs) for IV administration of rhu-pGSN in Sprague Dawley rats are:100 mg/kg/dose with 5 doses over 3 days, 100 mg/kg/dose daily for 7 days, and 25 mg/kg/dose daily for 14 days. The NOAEL for IV administration of rhu-pGSN in cynomolgus monkeys is 20 mg/kg/dose daily for 14 days.

Test article-related effects observed following repeat aerosol administration (3-25 mg/day) of rhu-pGSN under conditions of continuous pulmonary exposure were limited to minimal microscopic changes in the lungs of monkeys. The microscopic change is attributed to an immunogenic response as evidenced by the detection of antibodies to rhu-pGSN. The antibodies appeared to be directed against the human-specific epitopes of intact rhu-pGSN and caused no clinical or pathological signs of serum sickness. Thus, the immunogenic reaction to rhu-pGSN in monkeys appears to be a species-specific response and, therefore, is not considered to be clinically relevant. No eye (0.5 mg) or skin (2.5 mg/site) irritation was noted in rabbits after acute exposure to the product.

In summary, the preclinical profile of rhu-pGSN suggests that this drug will be well tolerated by humans and that the risk of respiratory or systemic toxicity is low.

##### 3.4.2. Clinical Experience

Rhu-pGSN has been previously evaluated in 5 clinical studies.

**Study 96-900** was a Phase 1, double-blind, randomized, controlled, within-subject, dose escalation study in 24 healthy volunteers conducted in the United Kingdom. Patients were dosed via nebulization up to a maximum dose of 32 mg given twice per day. Adverse events (AEs) were minor, and none were consistently attributed to treatment. No formation of antibodies to rhu-pGSN was observed.

**Study C96-901** was a randomized, double-blind, placebo-controlled, dose escalating, tolerability study of inhaled rhu-pGSN in 21 patients with cystic fibrosis conducted at one clinical site in Canada. Rhu-pGSN or placebo was administered via nebulization at 3.0 mg per day (Day 1 and 2), 10 mg per day (Day 3 and 4) and 25 mg per day (Day 5 to 9). Sixteen patients were randomized to rhu-pGSN and five patients to placebo (randomization ratio 3:1). Rhu-pGSN was well tolerated and no safety concerns were raised. No patients withdrew from the study. There was no negative effect on pulmonary function. There were no serious adverse events (SAEs), and no AEs were considered likely or definitely related to treatment.

**Study CBC-101** was a randomized, double-blind, placebo-controlled, ascending dose, infusion trial of the PK of rhu-pGSN in patients with decreased natural gelsolin levels. This Phase 1b/2a study was conducted in Hong Kong and enrolled patients admitted to the intensive care unit (ICU). Twenty-eight patients were enrolled; 21 patients received rhu-pGSN and 7 received placebo. The 4 cohorts were treated with ascending doses of rhu-pGSN via IV infusion according to the following scheme:

Cohort 1: Single infusion of 3 mg/kg rhu-pGSN (10 patients) or placebo (3 patients)

Cohort 2: Single infusion of 6 mg/kg rhu-pGSN (3 patients) or placebo (2 patients)

Cohort 3: Daily infusion of 6 mg/kg rhu-pGSN for 3 days (6 patients) or placebo (2 patients)

Cohort 4: Daily infusion of 6 mg/kg rhu-pGSN for 3 days (2 patients with severe multiple organ failure)

The dose escalation over the successive cohorts was supervised by an independent data and safety monitoring board (DSMB). The DSMB had to document the safety profile of the drug before allowing the study to proceed to a higher dose and/or exposure. No safety concerns associated with rhu-pGSN were raised by the DSMB. Seven patients died during the 3-month observation period (Table 2); none of the deaths were assessed as related to study treatment. No patient who received placebo died. The incidence of death in the rhu-pGSN-treated patients (7/21; 33%) is within the limits of expectation for patients admitted in ICU with similar diagnoses. However, the incidence of death was lower than expected in the placebo group, most probably due to imbalances in multiple baseline clinical prognostic factors in favor of the placebo group when compared to the rhu-pGSN group.

**Table 2: Summary of Deaths in Study CBC-101**

| Subject | Cohort | Age | pGSN mU/mL | Admission diagnosis[1] | VP | MV | Day of admission relative to infusion[2] | Day of death[3] |
|---|---|---|---|---|---|---|---|---|
| **G004** | 1 | 81-85 | 921 | Fecal peritonitis, emergency rectosigmoidectomy, renal insufficiency | Yes | Yes | -10 | 5 |
| **G006** | 1 | 66-70 | 1892 | Pneumonia with hepatitis | No | Yes | -10 | 3 |
| **G012** | 1 | 76-80 | 524 | Sigmoid perforation, emergency rectosigmoidectomy | No | No | -6 | 26 |
| **G015** | 2 | 76-80 | 1161 | Pneumonia, COPD | Yes | Yes | -15 | 21 |

| Subject | Cohort | Age | pGSN mU/mL | Admission diagnosis[1] | VP | MV | Day of admission relative to infusion[2] | Day of death[3] |
|---|---|---|---|---|---|---|---|---|
| **G017** | 2 | 76-80 | 963 | Peritonitis with terminal cholangiocarcinoma | No | No | -10 | 16 |
| **G018** | 2 | 81-85 | 1128 | Pneumonia with renal insufficiency | Yes | Yes | -10 | 66 |
| **G401** | 4 | 81-85 | 780 | Pneumonia with renal insufficiency | Yes | Yes | -4 | 7 |

COPD=chronic obstructive pulmonary disease; ICU=intensive care unit; MV=mechanical ventilation; pGSN=plasma gelsolin; VP=vasopressor.
[1] Diagnosis established at ICU admission
[2] Represents the day of hospitalization (irrespective of ICU or not) relative to day of infusion
[3] Day of death relative to first infusion

**Study BTI-201** was a Phase 1b/2a, double-blind, placebo-controlled, single and multiple ascending dose escalation study to evaluate the safety, PK, and pharmacodynamics of rhu-pGSN added to standard of care (SOC) in non-ICU patients hospitalized for mild community-acquired pneumonia (CAP). Eligible subjects were randomized 3:1 to receive adjunctive IV rhu-pGSN or placebo. Thirty-three subjects were treated: 8 received a single dose of rhu-pGSN 6 mg/kg, and 25 received a daily rhu-pGSN dose of 6, 12, or 24 mg/kg over 3 consecutive days (Tannous et al., 2020).

Overall, AEs were mild in both treatment groups irrespective of dose. AEs were reported more frequently in the lowest rhu-pGSN dose cohorts and the less frequently in the highest rhu-pGSN dose cohorts. Treatment-emergent adverse events (TEAEs) that were reported in more than 1 subject in the rhu-pGSN groups were: nausea (11.1% rhu-pGSN; 0% placebo) and blood pressure increased (11.1% rhu-pGSN; 0% placebo). No infusion-related reactions (IRRs) were reported. Of the subjects treated with rhu-pGSN, no subject experienced a TEAE assessed as drug-related, and no subject discontinued due to a TEAE.

Two subjects experienced SAEs: 1 SAE was reported in 1 subject who received a single dose of rhu-pGSN, and 2 SAEs were reported in 1 subject who received placebo in the multiple ascending dose phase. One subject who received a single dose of rhu-pGSN had an SAE of pneumonia (Grade 5, not related) after being withdrawn from the study; and 1 subject who received placebo had SAEs of pneumonia (Grade 4, not related) and pulmonary embolism (Grade 5, not related).

Pharmacokinetic analyses showed that the median rhu-pGSN half-life exceeded 17 hours with all dosing regimens. Overall, rhu-pGSN was well tolerated in patients admitted to non-ICU beds with CAP.

**Study BTI-202** was a Phase 2 randomized, double-blind, placebo-controlled study to evaluate the efficacy and safety of rhu-pGSN added to SOC for the treatment of subjects hospitalized with severe COVID-19 pneumonia (NCT04358406) (DiNubile et al., 2022). The study was conducted in Spain and Romania. Sixty-four subjects with severe COVID-19 pneumonia were randomized 1:1 to receive rhu-pGSN or placebo intravenously in addition to SOC. Standard of care was left to the judgment of the caregivers without restrictions. Three doses of rhu-pGSN 12 mg/kg based on actual body weight were administered at 0, 12, and 36 hours. Of the 61 treated subjects, 54 completed the study, 4 died, and 3 were lost to follow-up. No subject discontinued due to an AE. The primary efficacy endpoint, the proportion of subjects alive without respiratory, hemodynamic, or renal support on Day 14, did not differ between treatment arms: 25 of 30 rhu-pGSN recipients (83.3%) and 27 of 31 placebo recipients (87.1%). Adverse events occurred with similar frequencies in both treatment groups. Most AEs were mild to moderate in severity and related to COVID-19 pneumonia. Serious adverse events were numerically fewer in the rhu-pGSN group than placebo group and all were assessed as unrelated to study drug. No pattern of AEs was discernable in either treatment arm. Two subjects in each group died during the 90-day study, but no deaths were attributable to study drug.

#### 3.5. Rationale for Study and Starting Dose

Given that pGSN is a naturally occurring plasma protein, the risk of drug toxicity related to rhu-pGSN infusion is expected to be low. In healthy primates dosed for 14 days, drug levels were similar to those obtained in human patients with pneumonia after administration of 12 mg/kg without evident toxicities. The doses given to humans have reached 24 mg/kg for 3 consecutive days. No distinctive safety signal was seen in the 2 small BioAegis studies to date; in fact, SAEs were numerically fewer in rhu-pGSN recipients compared to saline-placebo control subjects. However, in Study CBC-101 evaluating sick patients in the ICU administered a previous formulation of rhu-pGSN, 7 of 21 rhu-pGSN and 0 of 7 placebo recipients died (Table 2). None of the deaths were judged to be related to study drug and were all attributed to the serious underlying conditions of the participants. A post-hoc analysis indicated the rhu-pGSN group had higher acuity scores than the placebo group. All but one subject who died in Study CBC-101 were in the single-dose rhu-pGSN dosing groups.

Efficacy has only been demonstrated in animal models. It is anticipated that in sick patients, pGSN levels would be lower than normal at presentation and that rhu-pGSN administered IV would be consumed more rapidly than in healthy subjects. As such, efficacy in sick patients may require doses higher than used in the earlier clinical trials. Based on the previous human experience, 3 daily doses of 6, 12, and 24 mg/kg of rhu-pGSN were well tolerated in patients with CAP. An accelerated dosing schedule where rhu-pGSN was administered to patients hospitalized with COVID-19 pneumonia as 12 mg/kg at time 0, 12, and 36 hours was also generally well tolerated.

Before treating a larger number of critically ill patients to establish proof of concept in subjects with ALI/ARDS, the Sponsor aims to study the safety, tolerability, and PK of IV rhu-pGSN administered to healthy subjects. The current study is designed to primarily evaluate the safety and tolerability of sequential ascending doses of rhu-pGSN at 6, 12, 18, and 24 mg/kg of body weight, each given 5 times at 0, 12, 36, 60, and 84 hours.

A risk analysis of the immunogenicity potential of rhu-pGSN has been conducted. Overall, the risk assessment based upon product, process, and subject factors indicates a low potential for generation of an immune response. Anti-pGSN antibodies could in theory be clinically meaningful. In the current study, specimens will be collected for ADA measurements at the first and last study visit.

### 4. STUDY OBJECTIVES AND ENDPOINTS

#### 4.1. Objectives

##### 4.1.1. Primary Objective

- To evaluate the safety and tolerability of 4 sequentially ascending doses of rhu-pGSN at 6 mg/kg, 12 mg/kg, 18 mg/kg, and 24 mg/kg of actual body weight administered by intravenous (IV) infusion for 5 doses at 0, 12, 36, 60, and 84 hours to healthy subjects

##### 4.1.2. Secondary Objectives

- To characterize the pharmacokinetic (PK) profile of rhu-pGSN during administration of multiple (5) doses of rhu-pGSN to healthy subjects assigned to 4 dose levels (6, 12, 18, or 24 mg/kg)
- To investigate the development of anti-pGSN antibodies following administration of multiple (5) doses of rhu-pGSN administered to healthy subjects assigned to 4 dose levels (6, 12, 18, or 24 mg/kg)

#### 4.2. Endpoints

##### 4.2.1. Primary Endpoint

- Incidence and severity of AEs regardless of causality graded according to the Toxicity Grading Scale for Healthy Adult and Adolescent Volunteers Enrolled in Preventive Vaccine Clinical Trials

##### 4.2.2. Secondary Endpoints

- PK for IV rhu-pGSN including, but not limited to: $C_{max}$, $T_{max}$, $T_{1/2}$, $AUC_{0-8}$, and additionally after Dose #5, $AUC_{0-24}$, and $AUC_{inf}$ (levels measured on Day 1 within 30 minutes predose, and at 15 minutes and 1, 2, 4, 8, and 12 hours after Dose #1 and on Days 1 and 2 and on Days 4 and 5 within 30 minutes predose, and at 15 minutes and 1, 2, 4, 8, 12, and 24 hours after Doses #2 and #5)
- Presence of anti-rhu-pGSN antibodies at Day 28

### 5. INVESTIGATIONAL PLAN

#### 5.1. Overall Study Design

Study BTI-101 is a Phase 1, randomized, double-blind, placebo-controlled, dose escalation, parallel design study to evaluate the safety, tolerability, and PK of IV rhu-pGSN or saline placebo administered as 5 doses each of 6, 12, 18, or 24 mg/kg of body weight. Each of 4 dosing cohorts will include 8 subjects randomized 3:1 rhu-pGSN:placebo (6 rhu-pGSN subjects:2 placebo subjects). Subjects will be healthy adults 18-50 years of age.

Doses will be administered at 0 hours (Day 1), 12 hours (Day 1), 36 hours (Day 2), 60 hours (Day 3), and 84 hours (Day 4). Subjects will be kept in-house until after the last blood sample is taken on Day 5. Subjects will return for follow-up 7 days after the initiation of therapy (Day 7) and on Day 28 for the End-of-Study (EOS) Visit. After each cohort has completed the Day 7 visit, review of the safety results (including labs) will be conducted (and unblinded where appropriate) before the initiation of the next higher dose cohort.

To assess safety and tolerability, subjects will undergo physical examinations (including vital sign measurements), AE assessments, concomitant medication assessments, and safety laboratory testing. Blood samples will be collected for analysis of pGSN levels and antibodies against pGSN. See Table 4 for a schedule of all study events.

#### 5.2. Safety Review Committee

The safety data will be reviewed by a Safety Review Committee (SRC) comprised of at least 1 expert clinician, the Medical Monitor at the contract research organization, and the Sponsor's Chief Medical Officer.

Adverse events (AEs) will be categorized as "serious" according to the judgement of the site investigator as to whether the definition of SAE (section 9.1.2) is met and then reviewed and adjudicated by the SRC. The "severity" of an AE will be graded according to the Toxicity Grading Scale for Healthy Adult and Adolescent Volunteers Enrolled in Preventive Vaccine Clinical Trials. An individual patient will be discontinued permanently from study treatment if they experience any serious adverse event (SAE) or a severity Grade 3 or greater AE (including infusion reactions). Further study treatment and enrollment will be paused pending review by the SRC if there are two Grade 3 AEs, one Grade 4 AE, or one SAE. The study will only be resumed if the SRC unanimously judges the AEs as unrelated to study treatment and procedures after consultation with regulatory and outside experts as needed.

The SRC will review safety data (including lab tests) through the Day 7 visit for each dosing arm to determine if it is safe to initiate the next cohort according to the safety criteria for stopping doses (Section 5.3).

#### 5.3. Safety Criteria for Stopping Doses

Dosing of rhu-pGSN may be stopped during the study at the discretion of the SRC. All AEs occurring during this healthy-volunteer study will be presumed to be related to study drug unless occurring before the study therapy is initiated pending review by the SRC.

An individual patient will be discontinued permanently from study treatment if they experience any serious adverse event (SAE) or a severity Grade 3 or greater AE (including infusion-related reactions) regardless of causality.

Further study treatment and enrollment will be paused pending review by the SRC if any patient dies or there are two Grade 3 AEs, one Grade 4 AE, or one SAE. The study will only be resumed if the SRC unanimously judges the death or AEs as unrelated to study treatment and procedures after consultation with regulatory and outside experts as needed.

##### 5.3.1. Management of rhu-pGSN Infusion-Related Reactions

In general, doses should only be temporarily interrupted for Grade 1 or 2 AEs, but treatment to control symptoms should be provided, if applicable. The infusion can be restarted at a slower rate once symptoms have dissipated, If symptoms recur, the infusion will be permanently discontinued. If a subject develops a Grade 3-4 IRR:

- The infusion should be stopped immediately, and the infusion tubing should be disconnected from the subject.
- The subject should receive appropriate treatment with an antihistamine and/or acetaminophen (paracetamol) and/or methylprednisolone (or equivalent) and, if necessary, further medications (i.e., epinephrine, bronchodilator).
- The subject may not receive further infusions of study therapy.

#### 5.4. Duration of Subject Participation

The duration of subject participation includes screening for up to 6 weeks, study treatment for 4 days, and safety follow-up through 28 days post-first dose for a maximum duration of subject participation of 10 weeks.

#### 5.5. End of Study Definition

End of study (EOS) will be defined as the date the last subject completes the EOS Visit. The Sponsor will notify all applicable regulatory agencies in accordance with local requirements when the study has ended.

#### 5.6. Criteria for Treatment Discontinuation

Subjects are free to discontinue their participation in the study at any time and without prejudice to further treatment. The Investigator must withdraw any subject from the study if that subject requests to be withdrawn, or if it is determined that continuing in the study would result in a significant safety risk to the subject.

The subject's participation in this study may be discontinued for the following reasons:

- Any SAE
- Grade 3 AEs or Grade 4 AE
- Subject withdrew consent
- Subject is unwilling or unable to continue the study or is lost to follow up

- Subject is non-compliant with study procedures/study protocol
- Investigator decides that withdrawal from the study is in the best interest of the subject
- Any clinically significant change in subject's medical condition (at the discretion of the Investigator)
- Sponsor decision to end the study

If a subject withdraws from the study prematurely, assessments scheduled for the EOS Visit should be performed as soon as possible. If a subject refuses further assessment, the subject should be contacted for safety evaluations (AE/concomitant medications/potential pregnancy) approximately 28 days after study withdrawal.

If such withdrawal occurs, or if the subject refuses to participate in the EOS Visit, the Investigator must determine the primary reason for a subject's withdrawal from the study and record the information on the electronic case report form (eCRF). If the reason for withdrawal is an AE, monitoring should continue until the outcome is evident. The specific event or test result(s) must be recorded in the eCRF. At the discretion of the Sponsor, subjects may also be removed from the study (Section 5.4).

It should be clearly documented in the source data whether a subject withdrew his/her consent and will not enter the follow-up phase, or if a subject withdrew his/her consent for study drug treatment but will continue further participation in the study.

#### 5.7. Criteria for Study Pausing or Termination

The Sponsor reserves the right to discontinue the study at any time for any reason. Such reasons may be any of, but not limited to, the following:

- Occurrence of AEs unknown to date in respect to their nature, severity, and duration, or the unexpected incidence of known AEs
- Any SAE pending review by the Safety Review Committee (SRC)
- Two Grade 3 or one Grade 4 AE pending review by the Safety Review Committee (SRC)
- Any death pending review by the Safety Review Committee (SRC)
- Medical or ethical reasons affecting the continued performance of the study

If the study is prematurely terminated, the Investigator is to promptly inform the study subjects and Independent Ethics Committee (IEC) and should ensure appropriate follow up for the subjects. All procedures and requirements pertaining to the archiving of study documents should be followed. All other study materials (e.g., study drug, etc.) must be destroyed or returned to the Sponsor.

### 6. STUDY POPULATION

#### 6.1. Number of Subjects

A total of 32 (6 rhu-pGSN and 2 placebo recipients per dose cohort x 4 dosing levels).

#### 6.2. Subject Inclusion Criteria

1. Healthy male or female adults 18 to 55 years of age without chronic or active acute medical conditions
2. Informed consent obtained from subject
3. Weight ≤100 kg and body mass index (BMI) <30 kg/$m^2$
4. Willingness during the course of the study, starting at screening and for at least 3 months after their final study treatment:
   a) Female subjects of childbearing potential must agree to use 2 medically accepted/Food and Drug Association (FDA)-approved birth control methods
   b) Male subjects with a partner who might become pregnant must agree to use reliable forms of contraception (i.e., condom, vasectomy, abstinence), or an acceptable method of birth control must be used by the partner
   c) All subjects must agree not to donate sperm or eggs

#### 6.3. Subject Exclusion Criteria

1. Pregnant or lactating women
2. Acute illness during the month prior to screening
3. Circumstances that may require any medications (including prescription medication, over-the-counter medication, vitamins, or supplements) during the during the 5 inpatient days of the study other than acetaminophen
4. Hospitalization during the year prior to screening
5. History of cancer or treatment with systemic chemotherapy or radiation therapy at any time
6. Transplantation of hematopoietic or solid organs
7. History of diabetes mellitus; myocardial infarction, angina, or other cardiovascular disease; stroke or cerebrovascular disease; chronic obstructive pulmonary disease (COPD) or asthma; deep vein thrombosis (DVT)/pulmonary embolism (PE); liver or kidney disease; clinically significant psychiatric condition; or active or chronic infection
8. Receipt of blood products during the year prior to screening
9. Chronic mechanical ventilation or dialysis
10. Any clinically significant abnormalities in clinical chemistry, hematology, or urinalysis results as judged by the Investigator

11. Any clinically significant abnormalities of vital signs, EKG or physical examination findings as judged by the Investigator
12. Positive results for recreational drugs during screening
13. Any other condition deemed by the Investigator as possibly interfering with the conduct of the study

#### 6.4. Subjects or Partners of Subjects of Reproductive Potential

Pregnancy is an exclusion criterion and women of childbearing potential must not be considering getting pregnant during the study. Unless they have had a hysterectomy, female subjects must have a negative serum or urine pregnancy test within 36 hours prior to start of study drug. A serum or urine pregnancy test will be performed at the EOS Visit. A total of 2 acceptable methods of birth control must be used per couple.

Women of childbearing potential must practice 2 acceptable methods of birth control starting at screening and continuing at least 3 months after the last study treatment. Acceptable methods of birth control in this study include true sexual abstinence (i.e., completely refraining from heterosexual intercourse throughout the study), hormonal methods (e.g., “the pill,” hormone injections, implants), barrier methods (e.g., condom or diaphragm), intrauterine device (IUD or “coil”), or sex exclusively with sterilized partner(s). An accepted birth control method used by the male partner can count as one of the 2 acceptable methods of birth control for the woman.

Male subjects with a partner who might become pregnant must use reliable forms of contraception (i.e., condom, vasectomy, abstinence) during the study starting at screening and for at least 3 months after the last study treatment, and an acceptable method of birth control must be used by the female partner (i.e., oral contraceptive, IUD, hormonal implants, contraceptive injection, or a double-barrier method).

Subjects will be instructed to notify the Investigator if pregnancy is discovered either during or within 3 months of the last dose of study drug.

#### 6.5. Waivers of Inclusion/Exclusion Criteria

No waivers of these inclusion or exclusion criteria (Section 6.2 and Section 6.3) will be granted by the Investigator and/or the Sponsor or its designee for any subject enrolling into the study.

### 7. DESCRIPTION OF STUDY TREATMENT

#### 7.1. Description of Study Drug

Rhu-pGSN drug product is provided as lyophilized powder containing rhu-pGSN plus excipients. It is provided in 10-mL glass vials to be reconstituted to a volume of 5.3 mL with 4.9 mL of sterile water yielding a final concentration of 40 mg/mL of rhu-pGSN, 2% glycine, 4% trehalose, 0.5% arginine, and 0.1% poloxamer 188 in a 10-mM phosphate buffer, pH 7 (Table 3).

Rhu-pGSN is administered via IV push ≤20 mL/minute. A total of 5 doses of rhu-pGSN will be administered to participating subjects at 0, 12, 36, 60, and 84 hours (Dose 1 is administered at time 0; a window of ±2 hours will be allowed around dosing times for the 2$^{nd}$ and 3$^{rd}$ doses, and ±4 hours for the last 2 doses).

**Table 3: Investigational Product**

| | **Investigational Product** |
|---|---|
| **Product Name:** | rhu-pGSN |
| **Dosage Form:** | Powder for solution filled as a 5.3 mL fill in 10 mL glass vials |
| **Unit Dose** | rhu-pGSN reconstituted to a volume of 5.3 mL with 4.9 mL of sterile water |
| **Route of Administration** | Intravenous |
| **Physical Description** | Lyophilized powder |
| **Storage Conditions** | 2 to 8 °C |
| **Manufacturer** | Drug Product: Lyophilization Services of New England, Inc., 25 Commerce Drive, Bedford, NH 03110, USA |

Vials containing rhu-pGSN study drug will be labeled according to national regulations for investigational products.

#### 7.2. Description of Placebo

Placebo will be dosed at a weight-based volume of 0.025 mL/mg × dose in mg/kg × weight in kg of 0.9% saline solution via IV push (matching the volume of rhu-pGSN for a subject of that size) injected at ≤20 mL/minute though a standard 0.2 μ filter at 0, 12, 36, 60, and 84 hours (Dose 1 is administered at time 0; a window of ±2 hours will be allowed around dosing times for the 2$^{nd}$ and 3$^{rd}$ doses, and ±4 hours for the last 2 doses). The placebo requires no special manipulation.

#### 7.3. Preparation of Study Drug for Administration

The Investigator or designee will be responsible for administering the appropriate dose of IV rhu-pGSN to subjects. rhu-pGSN must be stored refrigerated at 2 to 8 °C in its original package in an appropriate storage facility accessible only to the pharmacist(s), the Investigator, or a duly designated person.

The unblinded pharmacist will determine the dose (mg) of rhu-pGSN, calculate the volume of rhu-pGSN solution needed, and reconstitute lyophilized study drug with sterile water.

The individual rhu-pGSN infusion will be prepared under aseptic conditions and administered at the study site according to the directions of the Sponsor, which will be provided in a Pharmacy Manual. Any solution remaining in the vial must be discarded. After dilution for infusion, administration of rhu-pGSN should be initiated as soon as possible but no later than 6 hours. Maximum allowed storage times and conditions will be detailed in the Pharmacy Manual.

#### 7.4. Study Drug Administration

rhu-pGSN is administered via IV push ≤20 mL/minute. A total of 5 doses of rhu-pGSN will be administered to participating subjects at 0, 12, 36, 60, and 84 hours. Dose 1 is time 0; a window of ±2 hours will be allowed around dosing times for the 2$^{nd}$ and 3$^{rd}$ doses, and ±4 hours for the last 2 doses.

Subjects are to be monitored for administration site reactions during the infusion and for 1 hour after completion of each dose. Infusion site reactions will be recorded as AEs using the appropriate coding terms on the eCRF.

#### 7.5. Subject Monitoring During rhu-pGSN Infusion

Vital signs should be measured as outlined in Section 8.1.3. All supportive measures consistent with optimal patient care will be provided throughout the study according to institution standards.

Precautions for anaphylaxis should be observed during rhu-pGSN administration. Emergency resuscitation equipment and medications should be readily available. Additional supportive measures should also be available and may include, but are not limited to, epinephrine, antihistamines, corticosteroids, IV fluids, vasopressors, oxygen, bronchodilators, diphenhydramine, and acetaminophen (paracetamol).

#### 7.6. Shipment of Study Drug

Prior to study treatment, study medications will be supplied to the clinical trial site's pharmacy by the Sponsor or its designee.

Shipment of study drug supplies for the study will be accompanied by a shipment form describing the contents of the shipment, drug information, and other appropriate documentation. The shipment form will assist in maintaining current and accurate inventory records.

#### 7.7. Receipt and Storage of Study Drug

All study supplies should arrive at the pharmacy in sufficient quantity and in time to enable dosing as scheduled. The Investigator must ensure the acknowledgement of receipt of the clinical trial material (i.e., study drug and placebo) at the site, including that the material was received in good condition.

The Sponsor or its designee must notify the Investigator/study staff prior to dispatch of drug supplies, with the anticipated date of their arrival, addressed to the site's pharmacy.

The investigational drug will be stored in the pharmacy, refrigerated at 2 to 8 ºC. The Sponsor should be notified for any deviation from the storage conditions.

#### 7.8. Accountability, Handling, and Disposal of Study Drug

The study site must maintain accurate records documenting dates and amount of study drug received. The trial site's pharmacy will be responsible for ensuring the supervision of the storage and allocation of these supplies. When a shipment is received, the pharmacist will verify the quantities received and the accompanying documentation and will provide acknowledgment of receipt.

Accountability logs will be provided to assist the pharmacist in maintaining current and accurate inventory records covering receipt, dispensing, and disposition of the study drug. An unblinded study monitor will examine inventory during the study. Accountability records must be readily available and may be subject to inspection by regulatory authorities or independent auditors at any time.

Drug administration will be recorded in source documents and in the eCRFs.

At the end of the study, delivery records of study drug will be reconciled with used / unused supplies. A disposition form will be completed to verify that all used, unused or partially used supplies have been returned or destroyed following appropriate accountability review by the monitor. One copy of all accountability records and the disposition form will be retained by the Investigator for the study files.

#### 7.9. Concomitant Medications

Medications (i.e., prescriptions, over-the-counter medications, vitamins, or supplements) taken in the 30 days prior to study enrollment should be recorded and include the generic name (if possible). If known, document the start/stop date of each medication in the subject's file and in the eCRF.

During the study, no medications except acetaminophen are permitted.

#### 7.10. Treatment Compliance

The study drug will be administered by personnel at the study site to ensure compliance.

#### 7.11. Randomization and Blinding

Following screening, subjects qualified for study entry will be randomized to receive rhu-pGSN or placebo during the treatment period. All eligible subjects will be assigned a randomization number.

The investigational site team and the subject will be kept blinded to the treatment allocation of each participant. Only the designated pharmacist(s) will be unblinded to the treatment allocation. The unblinded pharmacist will randomly assign a treatment allocation.

##### 7.11.1. Unblinding

There is no antidote for rhu-pGSN. Unblinding should only be performed if knowledge of the treatment assignment will change the planned management of a medical condition or continuation of the trial. If possible, prior to unblinding, the need to unblind should be discussed with the Medical Monitor and Sponsor's Chief Medical Officer; the data safety committee should be involved re: decisions about unblinding that affect study pausing or termination if a short delay would not compromise medical care. However, this should not delay unblinding if the Investigator believes it is necessary. Each case of unblinding will be documented and documentation will be stored separately by the unblinded pharmacist.

Subjects that are unblinded may be withdrawn from the study. The decision to withdraw a subject from the study because of unblinding should be discussed with the Sponsor. If the subject is withdrawn, the Investigator or designee must record the date and reason for withdrawal on the appropriate eCRF for that subject.

### 8. STUDY ASSESSMENTS

The procedures and assessments that will be conducted during this study are described in this section and summarized in the Schedule of Assessments (Table 4). Detailed instructions regarding all laboratory procedures, including collection and handling of samples, will be included in the study Laboratory Manual provided by the Sponsor or its designee.

Written informed consent must be granted by each subject prior to the initiation of any study procedure or assessment.

Study visits will occur daily for 5 days (inpatient), at 7 ± 1 day after Dose #1 (outpatient), and at 28 ± 3 days after Dose #1 (outpatient). Subjects will be kept in-house until after the last blood sample is taken on Day 5 and asked to return on Days 8 and 28.

**Table 4: Schedule of Assessments**

| | Outpatient | Inpatient | Inpatient | Inpatient | Inpatient | Inpatient | Discharge | Outpatient | Outpatient |
|---|---|---|---|---|---|---|---|---|---|
| **Visit** | **Screening** | **Admission** | **Baseline/ Randomization/ Treatment** | **Treatment/Follow-Up Days** | | | | | **EOS Visit** |
| **Study Day** | **-6 weeks to -1** | **-1** | **1** | **2** | **3** | **4** | **5** | **8 ± 1** | **28 ± 3** |
| Informed consent | X | | | | | | | | |
| Eligibility assessment | X | X | | | | | | | |
| Admission to clinical research unit | | X | | | | | | | |
| Demographics | X | | | | | | | | |
| Medical history | X | X (interim) | | | | | | | |
| Blood or urine pregnancy test[a] | X | X[b] | | | | | | | X |
| Physical exam (incl. height and weight) | X | X | | | | | | | |
| Vital signs[c] | X | X | X | X | X | X | X | X | |
| EKG (after semi-recumbent for at least 5 min) | X | X | XX (pre-and post-dose 1 | | | X (post-dose 5 | | X | X |
| Blood/urine for safety laboratory analyses[d] | X | X[b] | | X (post-dose) | | | | X | X |
| Adverse events[e] | | | X | X | X | X | X | X | X |
| Review of Systems/Symptoms[e] | X | X | X | X | X | X | X | X | X |

| | Outpatient | Inpatient | Inpatient | Inpatient | Inpatient | Inpatient | Discharge | Outpatient | Outpatient |
|---|---|---|---|---|---|---|---|---|---|
| **Visit** | **Screening** | **Admission** | **Baseline/ Randomization/ Treatment** | **Treatment/Follow-Up Days** | | | | | **EOS Visit** |
| **Study Day** | **-6 weeks to -1** | **-1** | **1** | **2** | **3** | **4** | **5** | **8 ± 1** | **28 ± 3** |
| Study drug administration[f] | | | X/X (0 and 12 hours) | X (36 hours) | X (60 hours) | X (84 hours) | | | |
| Blood sampling for pGSN levels[g] | | | X (for Doses #1 and #2) | X (for Dose #2) | | X (for Dose #5) | X (for Dose #5) | X | X |
| Anti-rhu-pGSN antibody sampling | | | X | | | | | | X |
| Concomitant medications | X | X | X | X | X | X | X | X | X |

Abbreviations: AEs=adverse events; ALT=alanine aminotransferase; aPTT=activated partial thromboplastin time; AST=aspartate aminotransferase; CBC=complete blood count; CPK=creatine phosphokinase; EOS=end of study; PK=pharmacokinetic; PT=prothrombin time

Days refer to Study Days, not calendar days.

[a] Perform a serum or urine pregnancy test for all female subjects (unless they have had a documented hysterectomy).

[b] If performed more than 36 hours before initiation of study drug, the test must be repeated within this time frame.

[c] Record blood pressure, pulse oximetry, oral or infrared temperature, respiration rate, and heart rate after semi-recumbent for at least 5 min. To be measured during infusions at the following time points: pre-infusion and every 15 minutes during the infusion, at the end of infusion until 30 minutes after the end of infusion.

[d] Safety labs include collection of blood for a CBC, PT, aPTT, creatinine, bilirubin, alkaline phosphatase, CPK, AST, and ALT levels and collection of urine for drug screening (e.g., amphetamines, benzodiazepines, cannabinoids, cocaine, alcohol, and opiates) and urinalysis (dipstick: glucose, protein, red blood cells, white blood cells). Drug screening need only be performed once on Day -1.

[e] Subjects will be interviewed to assess any possible AEs daily for 5 days, at 8 ± 1 day after Dose #1, and at 28 ± 3 days after Dose #1 (EOS Visit). Interviews can be done in person or by phone as mutually agreed by the Investigator and subject, but the EOS Visit should be done in person if possible so blood specimens can be collected.

[f] Study drug will be administered by IV infusion at time 0 (Day 1), 12 hours (Day 1), 36 hours (Day 2), 60 hours (Day 3), and 84 hours (Day 4). Dose 1 will be at time 0, and a window of ±1 hour will be allowed around dosing times for the 2nd and 3rd doses, and ±2 hours for the last 2 doses. Subjects are to be monitored for administration site reactions during an infusion and for 1 hour after its completion.

[g] Approximately 5 mL volume of blood will be drawn for measurement of pGSN levels on Day 1 within 15 minutes predose, and at 15 minutes and 1, 2, 4, 8, and 12 hours after Dose #1 and on Days 1 and 2 and on Days 4 and 5 within 15 minutes predose, and at 15 minutes and 1, 2, 4, 8, 12, and 24 hours after Dose #2 and Dose #5, respectively. PK levels are timed from the end of the infusion, Plasma will be obtained after centrifugation and promptly frozen at -20°C or below

(down to -80°C) for assay in the central lab. A window of ±15 minutes will be allowed around the scheduled times except for the samples collected at 15 minutes postdose which have a window of ±5 minutes. A single sample for endogenous pGSN levels will be collected anytime on Days 8 and 28.

#### 8.1. Safety Assessments

Prior to screening, all subjects must provide informed consent for study participation, per Section 12.5. The following assessments will be performed according to the time points in the Schedule of Assessments (Table 4).

##### 8.1.1. Pregnancy Test

Unless they have had a hysterectomy, female subjects are required to have a negative serum or urine pregnancy test at screening to participate in the study. If the screening test is performed more than 36 hours before initiation of study drug, the test must be repeated within this time frame.

##### 8.1.2. Demographic/Medical History

A review of demographic parameters, including age, gender, race, and ethnicity will be performed at screening.

Past and present medical history will be recorded. Any ongoing condition or signs and symptoms observed prior to the initiation of study treatment should be recorded as medical history.

##### 8.1.3. Vital Signs

Vital signs will include semi-recumbent blood pressure, oral temperature, respiration rate, and heart rate, as well as pulse oximetry. Vitals signs will be measured during infusions at the following time points: pre-infusion, every 15 minutes after the start of infusion, at the end of infusion, and 30 minutes after the end of infusion. EKGs need to be performed in the semi-recumbent position as indicated in the schedule-of-assessments table 4. Significant findings noticed after the start of study drug that meet the definition of an AE must be recorded in the eCRF.

##### 8.1.4. Physical Examination

All physical examinations, including measurement of body height and weight for calculation of BMI, will be performed by a study physician or designee. Height (and BMI) need only be measured at the screening and admission examinations. The initial body weight measured at enrollment will be used to calculate rhu-pGSN dosing for all doses (so that an individual subject will always receive the same dose). The physical examination includes skin, head, ears, eyes, nose, throat, heart, lungs, abdomen, and neurologic system. Additional examination may be performed as found relevant by the Investigator.

##### 8.1.5. Laboratory Assessments

Clinical labs will be done locally. Where indicated (pGSN levels and ADA assays), handling and shipping laboratory samples to a central laboratory will be outlined in the Laboratory Manual. If a laboratory screening assessment is performed more than 36 hours before initiation of study drug, the test must be repeated within this time frame. Drug testing is only needed at screening and admission.

Blood will be collected at the time points specified in Table 4 for safety laboratory evaluations including complete blood count [CBC], prothrombin time (PT), activated partial thromboplastin time (aPTT), creatinine, bilirubin, alkaline phosphatase, creatine phosphokinase [CPK], aspartate aminotransferase [AST], and alanine aminotransferase [ALT] levels.

In addition, urine will be collected at the time points specified in Table 4 for drug screening (e.g., amphetamines, benzodiazepines, cannabinoids, cocaine, and opiates) and urinalysis (dipstick: glucose, protein, red blood cells, white blood cells).

Abnormal laboratory test results considered clinically significant by the Investigator or that require treatment should be reported as AEs in the eCRF.

#### 8.2. Measurement of pGSN Levels and Pharmacokinetic Assessments

As indicated in Table 4, approximately 5 mL volume of blood will be drawn for measurement of pGSN levels for PK assessments:

- on Day 1: within 15 minutes before Dose #1, and then at 15 minutes and 1, 2, 4, 8, and 12 hours after Dose #1,
- on Days 1 and 2: within 15 minutes before Dose #2, and then at 15 minutes and 1, 2, 4, 8, 12, and 24 hours after Dose #2, and
- on Days 4 and 5: within 15 minutes before Dose #5, and then at 15 minutes and 1, 2, 4, 8, 12, and 24 hours after Dose #5.

The post-dose sample 12 hours after Dose #1 will coincide with the pre-dose sample before Dose #2, and a single specimen can be used for this timepoint. A window of ±15 minutes will be allowed around the scheduled times except for the samples collected at 15 minutes post-dose which have a window of ±5 minutes. Post-dose times are measured from the end of the infusion. A single sample will be collected anytime on Days 8 and 28.

Plasma will be obtained after centrifugation and promptly frozen between -20°C and -80°C for assay in the central lab for analysis of pGSN concentrations. The concentration-time data will be used for the determination of PK parameters: maximum concentration ($C_{max}$), time to maximum concentration ($T_{max}$), terminal half-life ($T_{1/2}$), area under the curve from time zero to 8 hours ($AUC_{0-8}$), area under the curve from time zero to 12 hours ($AUC_{0-12}$), and additionally after Dose #5, area under the curve from time zero to 24 hours ($AUC_{0-24}$) and time zero to infinity ($AUC_{inf}$).

### 9. ADVERSE EVENT MANAGEMENT

#### 9.1. Definition of Adverse Events

##### 9.1.1. Adverse Event (AE)

An AE is the development of an undesirable medical condition or the worsening of a pre-existing medical condition following or during exposure to a pharmaceutical product, whether or not considered causally related to the product.

##### 9.1.2. Serious Adverse Event (SAE)

A serious adverse event (SAE) is an AE occurring during any study phase (i.e., baseline, treatment, or follow-up), and at any dose of the investigational product, comparator, or placebo, that fulfills one or more of the following:

- Results in death
- It is immediately life-threatening
- It requires in-patient hospitalization or prolongation of existing hospitalization
- It results in persistent or significant disability or incapacity
- Results in a congenital abnormality or birth defect
- It is an important medical event that may jeopardize the subject or may require medical intervention to prevent one of the outcomes listed above.

All SAEs that occur after a subject has signed the consent form, including before or during treatment and within 28 days following the cessation of treatment, whether or not judged to be related to the study, must be recorded on forms provided by the Sponsor.

#### 9.2. Clarifications to Serious Adverse Event Reporting

Death is an outcome of an SAE and not an SAE in itself. When death is an outcome, report the event(s) resulting in death as the SAE term (e.g., "pulmonary embolism"). If the cause of death is unknown, report "Death, unknown cause" as the SAE term.

#### 9.3. Assessment of Causality

All AEs occurring during this healthy-volunteer study will be presumed to be related to study drug unless occurring before the study therapy is initiated pending review by the SRC. AEs in the placebo group, although possibly related to a study procedure, cannot be related to rhu-pGSN unless the subject was mistakenly given the wrong treatment.

#### 9.4. Assessment of Severity

The severity rating of an AE refers to its intensity. The severity of each AE will be graded according to the Toxicity Grading Scale for Healthy Adult and Adolescent Volunteers Enrolled in Preventive Vaccine Clinical Trials (https://www.fda.gov/regulatory-information/search-fda-guidancedocuments/toxicity-grading-scale-healthy-adult-and-adolescent-volunteers-enrolledpreventive-vaccine-clinical).

It is important to distinguish between serious and severe AEs. Severity is a measure of intensity whereas seriousness is defined by the criteria under Section 9.1.2. An AE of severe intensity may not be considered serious.

#### 9.5. Pregnancy or Drug Exposure during Pregnancy

If a subject becomes pregnant during the study, administration of study drug is to be discontinued immediately.

Pregnancies must be reported within 24 hours of the Investigator's knowledge using the Sponsor's pregnancy form.

Pregnancy in itself is not regarded as an AE or SAE unless there is a suspicion that an investigational product may have interfered with the effectiveness of a contraceptive medication.

The outcome of all pregnancies (spontaneous miscarriage, elective termination, normal birth or congenital abnormality) must be followed up and documented even if the subject was discontinued from the study.

All reports of congenital abnormalities/birth defects are SAEs. Spontaneous miscarriages should also be reported and handled as SAEs. Elective abortions without complications should not be handled as AEs.

#### 9.6. Laboratory Abnormalities

The severity of each laboratory AE will be graded according to the Toxicity Grading Scale for Healthy Adult and Adolescent Volunteers Enrolled in Preventive Vaccine Clinical Trials (https://www.fda.gov/regulatory-information/search-fda-guidancedocuments/toxicity-grading-scale-healthy-adult-and-adolescent-volunteers-enrolledpreventive-vaccine-clinical).

To the extent possible, all laboratory abnormalities observed during the course of the study will be included under a reported AE term describing a clinical syndrome (e.g., elevated blood urea nitrogen and creatinine in the setting of an AE of "renal failure"). In these cases (e.g., an AE of renal failure), the laboratory abnormality itself (e.g., elevated creatinine) does not need to be recorded as an AE.

If a laboratory abnormality cannot be reported as a clinical syndrome, AND if the laboratory abnormality results in a therapeutic intervention (i.e., concomitant medication or therapy), or is judged by the Investigator to be of other clinical relevance, then the laboratory abnormality should be reported as an AE.

Subjects experiencing AEs or clinically significant laboratory abnormalities will be assessed and appropriate evaluations performed until all parameters have returned to baseline levels or are consistent with the subject's then-current physical condition.

#### 9.7. Reporting Adverse Events

All AEs, serious and nonserious, will be fully documented on the appropriate eCRF. For each AE, the Investigator must provide its duration (start and end dates or ongoing), intensity, assessment of causality and whether specific action or therapy was required. All AEs occurring during this healthy-volunteer study will be presumed to be related to study drug unless occurring before the study therapy is initiated pending review by the SRC.

All AEs that occur from the signing of the informed consent form (ICF) until the first dose of study drug should be recorded on the AE eCRF page only if the event was related to a study procedure. All other AEs/findings prior to the first dose of study drug should be recorded as medical history on the applicable eCRF page. All AEs occurring from the first dose of study drug until 28 days after the last dose of study drug must be recorded on the AE eCRF.

All SAEs and Grade 4 AEs must be reported to the Sponsor within 24 hours of the Investigator's knowledge. All AEs occurring during this healthy-volunteer study will be presumed to be related to study drug unless occurring before the study therapy is initiated pending review by the SRC. This reporting should be done by faxing/emailing the completed SAE Report Form to the number provided on the SAE Report Form. After the EOS Visit, only new treatment-related SAEs need to be captured on the AE eCRF and reported to the Sponsor.

Investigators must follow subjects with AEs/SAEs until the event has resolved, the condition has stabilized, withdrawal of consent, the subject is lost to follow up or death OR until the EOS Visit, whichever occurs first. New and ongoing treatment-related SAEs should be followed beyond the EOS Visit. If the subject dies, this should be captured as the outcome of the AE unless no link between the AE and the subject death can be established, in which case the AE will be marked as ongoing and the death will be reported as a separate event.

If a subject is lost to follow up, this should be captured accordingly within the AE eCRF and on the follow up SAE report.

The Sponsor or designee is responsible for notifying the relevant regulatory authorities of applicable suspected unexpected serious adverse reactions (SUSARs) as individual notifications or through periodic line listings. It is the Principal Investigator's responsibility to notify the Institutional Review Board (IRB) or IEC of all SAEs that occur at his or her site. Investigators will also be notified of all unexpected, serious, and drug-related events (7/15 Day Safety Reports) that occur during the clinical trial. Each site is responsible for notifying its IRB or IEC of these additional SAEs.

### 10. STATISTICS

#### 10.1. General Overview

##### 10.1.1. Method of Assigning Subjects to Treatment Groups

For each dosing group, 8 subjects will be randomized in a 3:1 ratio to receive an equal volume of rhu-pGSN or saline placebo during the treatment period. See Section 7.11 for details regarding randomization.

##### 10.1.2. Data Collection and Analysis

Baseline characteristics, safety, PK, and development of ADAs will be summarized via summary statistics separately for each rhu-pGSN dose level; data for all placebo subjects will be pooled. Additional details will be provided in a separate Statistical Analysis Plan (SAP).

#### 10.2. Sample Size and Power Calculations

Prior studies of rhu-pGSN evaluated 3 doses of rhu-pGSN in subjects hospitalized with CAP or COVID-19. To expand our understanding of possible rhu-pGSN toxicities in the absence of underlying disease, 24 healthy subjects will be administered 5 doses of rhu-pGSN at 6, 12, 18, and 24 mg/kg of body weight (6 subjects per dose group) and 8 healthy subjects (2 subjects per dose group) will be given the same volume on a weight basis of a visually indistinguishable 0.9% saline placebo to provide additional data to inform the safety and tolerability of rhu-pGSN.

Table 5 presents the minimum sample size such that there is a 90% probability of observing at least 1 AE of a certain type if the TRUE underlying AE incidence is as specified.

**Table 5: Minimum Sample Size Calculations**

| Sample Size | TRUE Underlying AE Rate |
|---|---|
| 6 (each rhu-pGSN dose level) | 32% |
| 8 (pooled placebo) | 25% |
| 12 | 18% |
| 18 | 13% |
| 24 (pooled rhu-pGSN doses) | 10% |

With each sample size presented in Table 5, if zero AEs of a certain type (e.g., SAEs) are observed, one could be "90% confident" that the TRUE underlying incidence for that AE is at most the percentage indicated in Table 5. Thus with 24 healthy subjects, if no SAEs are observed, the probability is 90% that the true underlying incidence of SAEs in healthy subjects administered 5 doses of rhu-pGSN at 6, 12, 18, or 24 mg/kg of body weight is at most 10%.

#### 10.3. Analysis Populations

No data will be excluded from the summary statistics and analyses, and missing data will not be imputed. All subjects given at least part of 1 dose of study drug will be included in the intention-to- treat analysis (full analysis set). As a sensitivity analysis, a per-protocol analysis set excluding subjects who missed doses and/or randomly discontinued the study before the primary Day 7 Visit will be analyzed. The expectation is that all analysis sets will be almost identical.

#### 10.4. Criteria for Evaluation and Statistical Methods

##### 10.4.1. Safety

Descriptive summaries of the incidence of all AEs, drug-related AEs, SAEs, drug-related SAEs, AEs leading to discontinuation, and deaths will be provided. All AEs and drug-related AEs will be summarized by counts and proportions of subjects having an AE, each AE type, and an AE of each System Organ Class. Serious AEs will be summarized similarly. AEs leading to discontinuation of treatment or the study will be summarized. All reasons for any early discontinuations will be summarized.

Laboratory data will be summarized in relation to normal range values via counts and percentages of subjects below, within, and above the respective normal range, and by summary statistics (N, mean, median, standard deviation, minimum, maximum, 90% confidence intervals, etc.) for baseline, each observed time point, and change from baseline at each observed time point for continuous lab endpoints.

##### 10.4.2. Baseline Characteristics

Demographics and medical history characteristics will be summarized via counts and percentages of subjects for categorical variables, and by N, mean and standard deviation or median and interquartile range, as appropriate for the distribution of continuous variables.

##### 10.4.3. Pharmacokinetics and Immunogenicity

$C_{max}$, $T_{max}$, $T_{1/2}$, $AUC_{0\text{-}8}$, $AUC_{0\text{-}12}$, and additionally after Dose #5, $AUC_{0\text{-}24}$ and $AUC_{inf}$ will be calculated from the pGSN levels measured for Doses #1 and #5. Since the assay measures endogenous pGSN levels as well as exogenous recombinant pGSN, PK parameters will be calculated for values above baseline by subtracting predose pGSN level from each observed concentration. Tables and graphs will be provided.

Antibodies against rhu-pGSN will be assayed from frozen specimens to determine whether the investigational product induces an antibody response in recipients. The frequency of anti-pGSN antibodies with 95% confidence intervals will be provided for Day 1 (predose) and Day 28. If any subjects had antibodies at Day 1, the presence or absence of antibodies at Days 1 and 28 will be summarized in a 2 × 2 table (Day 1 vs. Day 28).

### 11. DATA RECORDING, RETENTION AND MONITORING

#### 11.1. Case Report Forms

Data will be collected using an electronic data capture (EDC) system at the clinical site. The Investigator or designee will record data specified in the protocol using eCRFs. Changes or corrections to eCRFs will be made by the Investigator or an authorized member of the study staff according to the policies and procedures at the site and the eCRF completion guidelines.

It is the responsibility of the Investigator to ensure eCRFs are complete and accurate. Following review and approval, the Investigator or designee will electronically sign and date the pages. This signature certifies that the Investigator has thoroughly reviewed and confirmed all data on the eCRF. Regardless of whether this responsibility has been delegated, the Investigator is personally responsible for the accuracy and completeness of all data included in the eCRF.

The Sponsor or designee will provide a portable document format (PDF) file of the eCRFs to the site for archiving after all data have been monitored and reconciled.

#### 11.2. Records Retention

Per Good Clinical Practice (GCP) guidelines regarding records retention, study documents are to be retained at the site until at least 2 years after the last approval of a marketing application in an International Council for Harmonization of Technical Requirements for Pharmaceuticals for Human Use (ICH) region or at least 2 years have elapsed since the formal discontinuation of clinical development of study drug. The Sponsor will notify the site of the date when study documentation may be destroyed.

#### 11.3. Data Monitoring

This study will be closely monitored by representatives of the Sponsor throughout its duration. Monitoring will include personal visits with the Investigator and study staff as well as appropriate communications by telephone, fax, mail, email or use of the EDC system, if applicable. It is the responsibility of the monitor to inspect eCRFs at regular intervals throughout the study to verify the completeness, accuracy and consistency of the data and to confirm adherence to the study protocol and to GCP guidelines. The Investigator agrees to cooperate with the monitor to ensure that any problems detected during the course of this study are resolved promptly. The Investigator and site will permit study-related monitoring, audits, IEC review and regulatory inspection, including direct access to source documents.

It is understood that study monitors and any other personnel authorized by the Sponsor and/or Sponsor representatives may contact and visit the Investigator and will be permitted to inspect all study records (including eCRFs and other pertinent data) on request, provided that subject confidentiality is maintained and that the inspection is conducted in accordance with local regulations.

Every effort will be made to maintain the anonymity and confidentiality of subjects during this study. However, because of the experimental nature of this treatment, the Investigator agrees to allow representatives of the Sponsor as well as authorized representatives of regulatory authorities to inspect the facilities used in the conduct of this study and to inspect, for purposes of verification, the hospital or clinic records of all subjects enrolled in the study.

#### 11.4. Quality Control and Quality Assurance

The study site may be audited by a quality assurance representative of the Sponsor or its designee for the purpose of monitoring any aspect of the study. The Investigator agrees to allow the monitor and/or auditor to inspect the drug storage area, study drug stocks, drug accountability records, patient charts and study source documents, and other records relative to study conduct.

The Investigator should contact the Sponsor or designee immediately if contacted by a regulatory agency about an inspection.

### 12. REGULATORY, ETHICAL AND LEGAL OBLIGATIONS

#### 12.1. Good Clinical Practice

The study will be performed in accordance with the protocol, guidelines for GCP established by the ICH, and applicable local regulatory requirements and laws.

#### 12.2. Independent Ethics Committee Approval

The Investigator must inform and obtain approval from the IEC for the conduct of the study at named sites, the protocol, informed consent documents and any other written information that will be provided to the subjects and any advertisements that will be used. Written approval must be obtained prior to recruitment of subjects into the study and shipment of study drug.

The Investigator is responsible for informing the IEC of any amendment to the protocol in accordance with local requirements. Amendments may be implemented only after a copy of the approval letter from the IEC has been transmitted to the Sponsor. Amendments that are intended to eliminate an apparent immediate hazard to subjects may be implemented prior to receiving Sponsor or IEC approval. However, in this case, approval must be obtained as soon as possible after implementation.

Per GCP guidelines, the Investigator will be responsible for ensuring that an annual update is provided to the IEC to facilitate continuing review of the study and that the IEC is informed about the end of the study. Copies of the update, subsequent approvals and final letter must be sent to the Sponsor.

#### 12.3. Regulatory Authority Approval

The study will be performed in accordance with regional regulatory requirements and will also meet all of the requirements of ICH GCP guidance. Amendments to the protocol will be submitted to the appropriate regulatory agency/agencies prior to implementation in accordance with applicable regulations.

#### 12.4. Other Required Approvals

In addition to IEC and regulatory authority approval, all other required approvals (e.g. approval from the local research and development board or scientific committee) will be obtained prior to recruitment of subjects into the study and shipment of study drug.

#### 12.5. Informed Consent

It is the responsibility of the Investigator (or designee) to obtain written informed consent from each subject after adequate explanation of the aims, methods and potential hazards of the study and before any study procedures are initiated. The subject should be given the opportunity to ask questions and allowed time to consider the information provided. Each subject should be given a copy of the informed consent document and associated materials. The original copy of the signed and dated informed consent document must be retained at the site and is subject to inspection by representatives of the Sponsor or regulatory authorities; the subject should be given a copy of the signed ICF.

Substantial changes to the study protocol may necessitate modifications to the informed consent document. If an amended informed consent document is issued during a subject's participation in the study, the subject is required to provide written informed consent using the updated consent form prior to continuing with study-related activities.

Subjects unable or unwilling to provide written informed consent will not be enrolled in the study.

#### 12.6. Subject Confidentiality

The Investigator must ensure that subjects' privacy is maintained. On the eCRF or other documents submitted to the Sponsor, subjects will be identified by a subject number or a subject number and initials only. Documents that are not submitted to the Sponsor (e.g., signed informed consent documents) should be kept in a strictly confidential file by the Principal Investigator.

The Investigator shall permit authorized representatives of the Sponsor, regulatory authorities and IECs to review the portion of the subject's medical record that is directly related to the study. As part of the required content of informed consent documents, the subject must be informed that his/her records will be reviewed in this manner.

#### 12.7. Disclosure of Information

Information concerning the study, patent applications, processes, scientific data or other pertinent information is confidential and remains the property of the Sponsor. The Principal Investigator may use this information for the purposes of the study only.

It is understood by the Principal Investigator that the Sponsor will use information obtained in this clinical study in connection with the clinical development program, and therefore may disclose it as required to other clinical Investigators and to regulatory authorities. In order to allow the use of the information derived from this clinical study, the Principal Investigator understands that he/she has an obligation to provide complete test results and all data obtained during this study to the Sponsor.

Verbal or written discussion of results prior to study completion and full reporting should only be undertaken with written consent from the Sponsor.

### 13. LIST OF REFERENCES

DiNubile MJ, Parra S, Castro-Salomo A, and Levinson, SL. Adjunctive recombinant human plasma gelsolin for severe coronavirus disease 2019 pneumonia. Open Forum Infect Dis 2022; 9(8):ofac357. doi: 10.1093/fid/ofac357. eCollection 2022 Aug.

Tannous A, Levinson SL, Bolognese J, et al. Safety and pharmacokinetics of recombinant human plasma gelsolin in patients hospitalized for non-severe community-acquired pneumonia. Antimicrob Agents Chemother 2020;64 (10). https://doi.org/10.1128/AAC.00579-20.
