## Supplemental file 2 - Inclusion and Exclusion Criteria and Supplementary Table 1-6 for "A Phase 1, Single-Center, Randomized, Double-Blind, Placebo-Controlled, Multiple-Dose Escalation Study for the Evaluation of the Safety, Tolerability, and Pharmacokinetics of Recombinant Human Plasma Gelsolin (rhu-pGSN) Following Intravenous Administration to Healthy Volunteers"

**SUPPLEMENTARY MATERIALS**

**Full Inclusion and Exclusion Criteria**

**Inclusion Criteria**

1. Healthy male or female adults 18 to 55 years of age without chronic or active acute medical conditions

2. Informed consent obtained from subject

3. Weight ≤100 kg and body mass index (BMI) <30 kg/m2

4. Willingness during the course of the study, starting at screening and for at least 3 months after their final study treatment:

a) Female subjects of childbearing potential agreed to use 2 medically accepted/Food and Drug Association (FDA)-approved birth control methods

b) Male subjects with a partner who might have become pregnant agreed to use reliable forms of contraception (i.e., condom, vasectomy, abstinence), and an acceptable method of birth control must have been used by the partner

c) All subjects agreed not to donate sperm or eggs

**Exclusion Criteria**

1. Pregnant or lactating women

2. Acute illness during the month prior to screening

3. Circumstances that required any medications (including prescription medication, over the-counter medication, vitamins, or supplements) during the inpatient days of the study other than acetaminophen

4. Hospitalization during the year prior to screening

5. History of cancer or treatment with systemic chemotherapy or radiation therapy at any time

6. Transplantation of hematopoietic or solid organs

7. History of diabetes mellitus; myocardial infarction, angina, or other cardiovascular disease; stroke or cerebrovascular disease; chronic obstructive pulmonary disease (COPD) or asthma; deep vein thrombosis (DVT)/pulmonary embolism; liver or kidney disease; psychiatric condition; or active or chronic infection

8. Receipt of blood products during the year prior to screening

9. Chronic mechanical ventilation or dialysis

10. Any clinically significant abnormalities in clinical chemistry, hematology, or urinalysis results as judged by the Investigator

11. Any clinically significant abnormalities of vital signs, electrocardiogram (EKG) or physical examination findings as judged by the Investigator

12. Positive results for recreational drugs during screening

Any other condition deemed by the Investigator as possibly interfering with the conduct of the study

**Supplementary Table 1. List of adverse events in the Safety Population**

| Treatment Dose or Placebo | AE Term | Toxicity Grading Scale | Relationship to Study Drug | Relationship to Study Procedure | Outcome | TEAE? |
|---|---|---|---|---|---|---|
| 6 mg/kg | Right Chest Wall Muscle Strain. | Moderate (Grade 2) | Definitely Unrelated | Definitely Unrelated | Resolved | Yes |
| 6 mg/kg | Lightheaded | Mild (Grade 1) | Definitely Unrelated | Definitely Related | Resolved | No |
| 12 mg/kg | UTI | Moderate (Grade 2) | Definitely Unrelated | Definitely Unrelated | Resolved | Yes |
| 12 mg/kg | Left Upper Extremity Ecchymosis | Mild (Grade 1) | Definitely Unrelated | Definitely Related | Resolved | Yes |
| 12 mg/kg | Muscle Fasciculations | Mild (Grade 1) | Definitely Unrelated | Definitely Unrelated | Resolved | Yes |
| 18 mg/kg | Constipation | Mild (Grade 1) | Possibly Related | Definitely Unrelated | Resolved | Yes |
| 18 mg/kg | Bilateral Anterior Forearm Rash[a] | Mild (Grade 1) | Probably Unrelated | Probably Unrelated | Resolved | Yes |
| 18 mg/kg | Bilateral Anterior Forearm Rash[a] | Mild (Grade 1) | Definitely Unrelated | Definitely Unrelated | Resolved | Yes |
| 18 mg/kg | Bilateral Knee Pain | Mild (Grade 1) | Definitely Unrelated | Definitely Unrelated | Resolved | Yes |
| 18 mg/kg | Fatigue | Mild (Grade 1) | Possibly Related | Definitely Unrelated | Resolved | Yes |
| 24 mg/kg | Headache | Moderate (Grade 2) | Definitely Unrelated | Definitely Unrelated | Resolved | No |
| 24 mg/kg | Dermatitis | Mild (Grade 1) | Definitely Unrelated | Definitely Unrelated | Resolved | Yes |
| 24 mg/kg | Epigastric Abdominal Discomfort | Mild (Grade 1) | Possibly Related | Definitely Unrelated | Resolved | Yes |
| Placebo | Headache | Mild (Grade 1) | Probably Unrelated | Definitely Unrelated | Resolved | Yes |

[a]The same AE occurred as two separate episodes in one subject and was therefore counted as two AEs. The first episode began 5 days after Dose 5 and resolved 9 days later, while the second episode began 25 days after Dose 5 and resolved after 5 days.

**Supplementary Table 2. Selected hematology parameters over time in the Safety Population**

| Parameter | rhu-pGSN | | | | Placebo |
|---|---|---|---|---|---|
| | 6 mg/kg N=6 | 12 mg/kg N=6 | 18 mg/kg N=6 | 24 mg/kg N=6 | N=8 |
| | Mean (standard deviation) | | | | |
| **Hematocrit (%)** | | | | | |
| Baseline | 40.77 (4.26) | 41.38 (3.21) | 41.97 (3.48) | 43.05 (2.78) | 43.53 (2.54) |
| Change from BL to: | | | | | |
| Day 2 | -0.13 (1.67) | 1.03 (1.50) | 1.70 (2.71) | -0.13 (1.05) | 1.86 (2.64) |
| Day 8 | -0.38 (1.77) | -1.90 (3.25) | -1.28 (1.01) | -1.35 (1.96) | -0.06 (2.50) |
| Day 28 | -0.08 (0.98) | -0.70 (1.71) | -1.07 (2.50) | -1.27 (1.96) | 0.20 (2.76) |
| **Hemoglobin (g/L)** | | | | | |
| Baseline | 132.8 (16.44) | 135.0 (14.41) | 136.8 (11.55) | 135.7 (12.47) | 140.1 (8.90) |
| Change from BL to: | | | | | |
| Day 2 | 1.0 (2.45) | 5.2 (3.54) | 4.5 (7.40) | 0.2 (4.31) | 6.0 (9.35) |
| Day 8 | -1.2 (5.00) | -5.0 (9.59) | -4.0 (3.52) | -4.0 (4.90) | 0.0 (9.04) |
| Day 28 | -2.2 (4.02) | -1.0 (5.40) | -3.8 (8.13) | -3.7 (4.80) | -0.3 (8.38) |
| **Lymphocytes ($10^9$/L)** | | | | | |
| Baseline | 2.20 (0.60) | 2.20 (1.16) | 1.81 (0.69) | 1.83 (0.29) | 1.74 (0.46) |
| Change from BL to: | | | | | |
| Day 2 | -0.02 (0.48) | -0.33 (0.71) | -0.28 (0.50) | -0.44 (0.24) | 0.08 (0.54) |
| Day 8 | -0.21 (0.23) | -0.22 (0.65) | -0.09 (0.17) | 0.06 (0.37) | 0.15 (0.30) |
| Day 28 | -0.16 (0.58) | 0.01 (0.51) | -0.19 (0.26) | -0.22 (0.29) | 0.01 (0.40) |
| **Platelets ($10^9$/L)** | | | | | |
| Baseline | 263.5 (50.06) | 283.3 (90.70) | 227.0 (53.21) | 303.8 (49.73) | 247.3 (53.43) |
| Change from BL to: | | | | | |
| Day 2 | -5.0 (13.48) | -8.2 (15.85) | -8.0 (13.37) | -0.7 (17.66) | 0.6 (18.44) |
| Day 8 | -6.2 (20.09) | -19.3 (69.71) | -6.5 (24.64) | 31.0 (19.15) | -3.3 (37.62) |
| Day 28 | -9.8 (18.35) | 3.8 (10.98) | -13.0 (15.91) | -6.8 (17.93) | 1.0 (36.27) |

**Supplementary Table 3. Summary of pGSN concentration (µg/mL) in each cohort pre-dose 1 in subjects who were part of both the PK and PP populations**

| | rhu-pGSN | | | | |
|---|---|---|---|---|---|
| Statistics | 6 mg/kg N=6 | 12 mg/kg N=6 | 18 mg/kg N=5 | 24 mg/kg N=6 | Placebo N=8 |
| Mean | 64.84 | 63.57 | 59.61 | 72.51 | 69.98 |
| SD | 13.22 | 19.65 | 8.79 | 11.12 | 11.22 |
| CV% | 20.39 | 30.92 | 14.75 | 15.34 | 16.04 |
| Geometric Mean | 63.60 | 61.19 | 59.08 | 71.77 | 69.21 |
| Geometric CV% | 22.42 | 30.56 | 15.20 | 16.03 | 16.01 |
| Min | 42.75 | 45.75 | 48.23 | 54.63 | 54.40 |
| Median | 65.69 | 57.79 | 60.34 | 71.80 | 69.66 |
| Max | 82.88 | 94.05 | 68.11 | 88.65 | 89.49 |

**Supplementary Table 4. Summary of baseline-adjusted pGSN concentration (μg/mL) after Dose 1 in subjects who were part of both the PK and PP populations**

| Timepoint | Statistics | rhu-pGSN 6 mg/kg N=6 | rhu-pGSN 12 mg/kg N=6 | rhu-pGSN 18 mg/kg N=5 | rhu-pGSN 24 mg/kg N=6 | Placebo N=8 |
|---|---|---|---|---|---|---|
| 15 Mins Post-Dose 1 | Mean | 147.52 | 339.84 | 399.42 | 642.01 | 2.48 |
| | SD | 25.15 | 43.77 | 47.70 | 147.06 | 4.02 |
| | CV% | 17.05 | 12.88 | 11.94 | 22.91 | 162.23 |
| | Geometric Mean | 145.67 | 337.62 | 397.17 | 627.63 | 5.93 |
| | Geometric CV% | 17.78 | 12.41 | 11.89 | 23.91 | 60.13 |
| | Min | 108.81 | 297.87 | 354.99 | 434.46 | 0 |
| | Median | 150.37 | 326.90 | 385.10 | 621.45 | 0 |
| | Max | 184.71 | 415.78 | 452.28 | 850.46 | 11.16 |
| 1 Hour Post-Dose 1 | Mean | 134.85 | 305.73 | 413.27 | 610.06 | 4.29 |
| | SD | 25.75 | 41.82 | 71.64 | 71.68 | 6.06 |
| | CV% | 19.10 | 13.68 | 17.34 | 11.75 | 141.34 |
| | Geometric Mean | 132.76 | 303.24 | 407.84 | 606.74 | 4.38 |
| | Geometric CV% | 19.72 | 14.26 | 18.80 | 11.31 | 178.03 |
| | Min | 103.04 | 247.78 | 302.05 | 541.96 | 0 |
| | Median | 136.01 | 320.76 | 408.78 | 587.94 | 2.36 |
| | Max | 166.85 | 344.32 | 478.65 | 736.88 | 17.69 |
| 2 Hours Post-Dose 1 | Mean | 135.99 | 285.81 | 346.81 | 582.53 | 2.62 |
| | SD | 28.82 | 51.54 | 73.97 | 79.80 | 4.96 |
| | CV% | 21.19 | 18.03 | 21.33 | 13.70 | 189.60 |
| | Geometric Mean | 133.27 | 281.94 | 340.91 | 578.03 | 5.32 |
| | Geometric CV% | 22.67 | 18.28 | 20.66 | 13.66 | 108.55 |
| | Min | 98.26 | 220.36 | 269.09 | 496.17 | 0 |
| | Median | 142.75 | 285.45 | 321.11 | 565.22 | 2.91 |
| | Max | 169.40 | 360.13 | 462.94 | 679.66 | 14.28 |

| | | | | | | |
|---|---|---|---|---|---|---|
| 4 Hours Post-Dose 1 | Mean | 121.81 | 252.80 | 358.38 | 459.00 | 2.80 |
| | SD | 30.53 | 30.72 | 84.03 | 65.59 | 5.17 |
| | CV% | 25.07 | 12.15 | 23.45 | 14.29 | 184.71 |
| | Geometric Mean | 117.89 | 251.29 | 350.41 | 455.09 | 3.76 |
| | Geometric CV% | 30.40 | 11.96 | 24.20 | 14.40 | 120.56 |
| | Min | 67.24 | 221.30 | 254.75 | 392.69 | 0 |
| | Median | 126.84 | 244.47 | 333.84 | 457.72 | 0.97 |
| | Max | 150.52 | 299.02 | 462.52 | 536.68 | 15.25 |
| 8 Hours Post-Dose 1 | Mean | 88.75 | 203.49 | 277.73 | 319.24 | 2.63 |
| | SD | 22.56 | 34.17 | 103.07 | 53.75 | 6.65 |
| | CV% | 25.42 | 16.79 | 37.11 | 16.84 | 252.55 |
| | Geometric Mean | 86.09 | 201.19 | 264.82 | 315.66 | 6.27 |
| | Geometric CV% | 28.54 | 16.51 | 34.23 | 16.40 | 326.13 |
| | Min | 53.47 | 164.13 | 190.81 | 255.63 | 0 |
| | Median | 92.15 | 197.39 | 254.25 | 314.06 | 0 |
| | Max | 116.96 | 258.24 | 451.68 | 411.89 | 18.98 |
| 12 Hours Post-Dose 1 | Mean | 80.73 | 166.36 | 187.93 | 258.86 | 3.19 |
| | SD | 10.59 | 29.71 | 23.59 | 56.69 | 4.61 |
| | CV% | 13.12 | 17.86 | 12.55 | 21.90 | 144.57 |
| | Geometric Mean | 80.11 | 164.39 | 186.75 | 253.54 | 4.92 |
| | Geometric CV% | 13.89 | 16.53 | 12.54 | 22.82 | 110.14 |
| | Min | 63.59 | 143.87 | 159.47 | 188.29 | 0 |
| | Median | 84.02 | 156.39 | 183.00 | 264.47 | 0.78 |
| | Max | 91.57 | 222.64 | 220.40 | 327.01 | 12.58 |

**Supplementary Table 5. Summary of baseline adjusted pGSN concentration (µg/mL) after Dose 2 in subjects who were part of both the PK and PP populations**

| Timepoint | Statistics | rhu-pGSN 6 mg/kg N=6 | rhu-pGSN 12 mg/kg N=6 | rhu-pGSN 18 mg/kg N=5 | rhu-pGSN 24 mg/kg N=6 | Placebo N=8 |
|---|---|---|---|---|---|---|
| 15 Mins Post-Dose 2 | Mean | 224.74 | 498.10 | 590.19 | 844.89 | 1.91 |
| | SD | 31.02 | 79.26 | 85.92 | 204.75 | 5.41 |
| | CV% | 13.80 | 15.91 | 14.56 | 24.23 | 282.84 |
| | Geometric Mean | 222.74 | 493.23 | 585.01 | 822.63 | 15.29 |
| | Geometric CV% | 15.15 | 15.20 | 15.07 | 26.39 | 15.29 |
| | Min | 167.32 | 404.86 | 491.99 | 533.85 | 0 |
| | Median | 228.74 | 484.54 | 631.28 | 846.36 | 0 |
| | Max | 253.48 | 644.60 | 671.26 | 1117.80 | 15.29 |
| 1 Hour Post-Dose 2 | Mean | 211.93 | 444.09 | 623.19 | 812.08 | 2.47 |
| | SD | 35.55 | 76.65 | 42.08 | 166.49 | 6.86 |
| | CV% | 16.77 | 17.26 | 6.75 | 20.50 | 278.15 |
| | Geometric Mean | 209.12 | 439.28 | 622.05 | 796.57 | 2.37 |
| | Geometric CV% | 18.69 | 15.75 | 6.77 | 22.38 | 8439.79 |
| | Min | 148.18 | 390.94 | 570.29 | 539.73 | 0 |
| | Median | 219.73 | 422.58 | 608.75 | 829.38 | 0 |
| | Max | 245.39 | 594.88 | 667.70 | 1039.55 | 19.44 |
| 2 Hours Post-Dose 2 | Mean | 202.07 | 432.00 | 567.52 | 752.02 | 3.90 |
| | SD | 24.86 | 69.60 | 50.70 | 148.36 | 6.21 |
| | CV% | 12.30 | 16.11 | 8.93 | 19.73 | 159.29 |
| | Geometric Mean | 200.83 | 427.52 | 565.68 | 738.22 | 1.74 |
| | Geometric CV% | 12.11 | 15.79 | 9.07 | 22.10 | 32803.63 |
| | Min | 175.92 | 351.62 | 504.74 | 495.85 | 0 |
| | Median | 199.89 | 419.26 | 580.85 | 771.93 | 0.01 |
| | Max | 242.16 | 545.91 | 620.54 | 923.27 | 17.00 |

| | | | | | | |
|---|---|---|---|---|---|---|
| 4 Hours Post-Dose 2 | Mean | 175.13 | 396.55 | 605.81 | 662.08 | 3.77 |
| | SD | 26.87 | 61.68 | 166.62 | 135.23 | 3.80 |
| | CV% | 15.34 | 15.56 | 27.50 | 20.43 | 100.65 |
| | Geometric Mean | 173.27 | 392.68 | 590.08 | 649.17 | 5.32 |
| | Geometric CV% | 16.47 | 15.35 | 25.14 | 22.76 | 68.03 |
| | Min | 130.84 | 313.18 | 477.17 | 433.13 | 0 |
| | Median | 182.45 | 391.92 | 556.79 | 677.79 | 3.45 |
| | Max | 199.14 | 503.47 | 890.63 | 827.95 | 9.13 |
| 8 Hours Post-Dose 2 | Mean | 149.65 | 310.72 | 467.50 | 480.36 | 3.39 |
| | SD | 22.49 | 34.66 | 152.92 | 81.90 | 4.44 |
| | CV% | 15.03 | 11.16 | 32.71 | 17.05 | 130.85 |
| | Geometric Mean | 148.24 | 309.02 | 451.21 | 473.66 | 5.90 |
| | Geometric CV% | 15.15 | 11.70 | 28.93 | 19.23 | 69.18 |
| | Min | 124.75 | 250.44 | 375.15 | 329.43 | 0 |
| | Median | 148.19 | 311.54 | 389.87 | 494.75 | 1.47 |
| | Max | 177.52 | 356.71 | 734.84 | 567.23 | 11.39 |
| 12 Hours Post-Dose 2 | Mean | 117.57 | 243.81 | 374.04 | 396.29 | 3.18 |
| | SD | 16.31 | 27.52 | 120.05 | 77.50 | 4.33 |
| | CV% | 13.87 | 11.29 | 32.10 | 19.56 | 136.09 |
| | Geometric Mean | 116.69 | 242.49 | 361.71 | 389.95 | 5.47 |
| | Geometric CV% | 13.32 | 11.48 | 27.94 | 19.98 | 70.51 |
| | Min | 99.06 | 207.57 | 304.71 | 283.80 | 0 |
| | Median | 113.79 | 245.99 | 322.49 | 387.12 | 1.30 |
| | Max | 147.16 | 276.99 | 587.54 | 522.11 | 12.14 |
| 24 Hours Post-Dose 2 | Mean | 72.92 | 142.73 | 217.96 | 204.47 | 1.64 |
| | SD | 19.31 | 24.52 | 74.60 | 50.05 | 4.61 |
| | CV% | 26.48 | 17.18 | 34.23 | 24.48 | 280.61 |
| | Geometric Mean | 70.66 | 141.08 | 209.08 | 199.00 | 1.09 |
| | Geometric CV% | 28.70 | 16.56 | 32.07 | 26.55 | 47466.00 |

| | | | | | | |
|---|---|---|---|---|---|---|
| | Min | 44.61 | 125.00 | 157.82 | 130.66 | 0 |
| | Median | 75.62 | 128.55 | 177.69 | 204.15 | 0 |
| | Max | 101.52 | 175.43 | 338.74 | 264.87 | 13.04 |

**Supplementary Table 6. Summary of baseline adjusted pGSN Concentration (µg/mL) after Dose 5 in subjects who were part of both the PK and PP populations**

| | | rhu-pGSN | | | | |
|---|---|---|---|---|---|---|
| **Timepoint** | **Statistics** | **6 mg/kg N=6** | **12 mg/kg N=6** | **18 mg/kg N=5** | **24 mg/kg N=6** | **Placebo N=8** |
| Pre-Dose 5 | Mean | 71.56 | 158.92 | 215.01 | 232.78 | 2.97 |
| | SD | 21.48 | 11.50 | 35.09 | 47.64 | 5.16 |
| | CV% | 30.01 | 7.24 | 16.32 | 20.47 | 173.46 |
| | Geometric Mean | 68.70 | 158.58 | 212.77 | 228.52 | 4.24 |
| | Geometric CV% | 32.90 | 7.20 | 16.23 | 21.61 | 110.40 |
| | Min | 41.33 | 145.39 | 177.38 | 165.37 | 0 |
| | Median | 72.99 | 158.37 | 214.55 | 236.99 | 0.99 |
| | Max | 102.32 | 175.99 | 265.01 | 288.13 | 15.19 |
| 15 Mins Post-Dose 5 | Mean | 203.27 | 471.99 | 713.73 | 915.23 | 2.25 |
| | SD | 25.21 | 39.68 | 140.74 | 142.28 | 3.73 |
| | CV% | 12.40 | 8.41 | 19.72 | 15.55 | 165.78 |
| | Geometric Mean | 201.88 | 470.61 | 703.83 | 905.89 | 5.28 |
| | Geometric CV% | 13.10 | 8.36 | 18.35 | 15.85 | 65.88 |
| | Min | 162.26 | 424.06 | 608.64 | 742.61 | 0 |
| | Median | 212.91 | 463.54 | 671.42 | 920.72 | 0 |
| | Max | 229.97 | 523.62 | 952.97 | 1068.97 | 10.43 |
| 1 Hour Post-Dose 5 | Mean | 196.83 | 465.08 | 589.77 | 904.59 | 1.23 |
| | SD | 27.41 | 90.50 | 33.21 | 50.22 | 2.72 |
| | CV% | 13.93 | 19.46 | 5.63 | 5.55 | 221.26 |
| | Geometric Mean | 195.05 | 457.75 | 589.02 | 903.42 | 4.06 |
| | Geometric CV% | 15.32 | 19.78 | 5.64 | 5.59 | 113.24 |
| | Min | 146.36 | 346.24 | 551.29 | 831.98 | 0 |
| | Median | 205.64 | 475.45 | 587.74 | 908.28 | 0 |
| | Max | 220.45 | 606.54 | 629.50 | 970.21 | 7.71 |

| | | | | | | |
|---|---|---|---|---|---|---|
| 2 Hours Post-Dose 5 | Mean | 199.48 | 432.58 | 590.03 | 837.83 | 1.08 |
| | SD | 35.78 | 62.02 | 71.51 | 99.84 | 3.06 |
| | CV% | 17.94 | 14.34 | 12.12 | 11.92 | 282.84 |
| | Geometric Mean | 196.85 | 428.77 | 586.55 | 832.60 | 8.65 |
| | Geometric CV% | 17.96 | 14.78 | 12.25 | 12.49 | 8.65 |
| | Min | 158.86 | 346.00 | 500.12 | 673.90 | 0 |
| | Median | 193.04 | 441.38 | 576.20 | 853.91 | 0 |
| | Max | 243.43 | 503.77 | 674.00 | 944.25 | 8.65 |
| 4 Hours Post-Dose 5 | Mean | 164.40 | 377.10 | 562.78 | 698.64 | 2.96 |
| | SD | 36.26 | 36.48 | 109.28 | 73.97 | 5.13 |
| | CV% | 22.05 | 9.68 | 19.42 | 10.59 | 173.31 |
| | Geometric Mean | 160.97 | 375.58 | 554.53 | 695.46 | 2.26 |
| | Geometric CV% | 23.12 | 9.96 | 19.28 | 10.43 | 1192.70 |
| | Min | 111.64 | 317.06 | 447.77 | 624.18 | 0 |
| | Median | 162.08 | 377.11 | 534.23 | 674.62 | 0.05 |
| | Max | 217.95 | 425.68 | 714.11 | 796.33 | 14.61 |
| 8 Hours Post-Dose 5 | Mean | 161.38 | 321.79 | 415.61 | 579.03 | 2.97 |
| | SD | 59.98 | 83.51 | 41.60 | 96.68 | 5.56 |
| | CV% | 37.17 | 25.95 | 10.01 | 16.70 | 187.35 |
| | Geometric Mean | 153.69 | 314.33 | 414.04 | 571.87 | 11.77 |
| | Geometric CV% | 33.93 | 22.97 | 9.64 | 17.76 | 19.17 |
| | Min | 101.60 | 254.84 | 377.06 | 421.74 | 0 |
| | Median | 144.97 | 300.40 | 400.46 | 571.18 | 0 |
| | Max | 276.06 | 486.83 | 485.09 | 703.65 | 13.47 |
| 12 Hours Post-Dose 5 | Mean | 144.06 | 241.32 | 340.89 | 469.81 | 2.47 |
| | SD | 61.37 | 61.60 | 11.61 | 51.02 | 5.93 |
| | CV% | 42.60 | 25.53 | 3.41 | 10.86 | 239.99 |
| | Geometric Mean | 135.37 | 235.64 | 340.74 | 467.54 | 6.92 |
| | Geometric CV% | 38.15 | 23.51 | 3.38 | 10.79 | 199.27 |

| | | | | | | |
|---|---|---|---|---|---|---|
| | Min | 92.98 | 177.94 | 329.12 | 410.43 | 0 |
| | Median | 124.62 | 223.07 | 336.65 | 455.69 | 0 |
| | Max | 260.94 | 358.55 | 357.95 | 537.69 | 16.95 |
| 24 Hours Post-Dose 5 | Mean | 125.93 | 156.87 | 228.02 | 269.42 | 2.66 |
| | SD | 58.01 | 19.42 | 29.52 | 26.21 | 5.44 |
| | CV% | 46.06 | 12.38 | 12.95 | 9.73 | 204.49 |
| | Geometric Mean | 115.78 | 155.91 | 226.57 | 268.37 | 4.86 |
| | Geometric CV% | 46.76 | 12.44 | 12.50 | 9.62 | 140.36 |
| | Min | 64.70 | 137.32 | 196.62 | 241.74 | 0 |
| | Median | 104.47 | 155.31 | 220.53 | 262.61 | 0 |
| | Max | 219.28 | 178.65 | 276.29 | 302.25 | 15.74 |
| Day 8 | Mean | 36.05 | 40.47 | 46.40 | 46.90 | 6.14 |
| | SD | 22.99 | 16.18 | 23.63 | 24.19 | 6.71 |
| | CV% | 63.77 | 39.98 | 50.92 | 51.57 | 109.31 |
| | Geometric Mean | 26.31 | 37.70 | 42.06 | 39.73 | 6.26 |
| | Geometric CV% | 145.49 | 44.32 | 51.93 | 80.84 | 95.85 |
| | Min | 3.49 | 18.83 | 24.39 | 11.01 | 0 |
| | Median | 35.69 | 39.74 | 36.58 | 48.37 | 3.54 |
| | Max | 70.44 | 67.34 | 82.46 | 76.38 | 19.57 |
| Day 28 | Mean | 19.67 | 8.37 | 18.47 | 9.30 | 4.17 |
| | SD | 16.29 | 9.53 | 7.68 | 10.72 | 6.05 |
| | CV% | 82.79 | 113.80 | 41.57 | 115.30 | 145.05 |
| | Geometric Mean | 18.40 | 2.12 | 17.13 | 6.87 | 6.54 |
| | Geometric CV% | 114.88 | 2526.80 | 46.49 | 170.53 | 102.09 |
| | Min | 0 | 0.03 | 10.47 | 0 | 0 |
| | Median | 20.14 | 5.24 | 18.70 | 4.64 | 1.17 |
| | Max | 43.89 | 23.50 | 27.72 | 27.10 | 16.63 |
